# MEGA: Machine Learning-Enhanced Graph Analytics for Infodemic Risk Management

**DOI:** 10.1101/2020.10.24.20215061

**Authors:** Ching Nam Hang, Pei-Duo Yu, Siya Chen, Chee Wei Tan, Guanrong Chen

## Abstract

The COVID-19 pandemic brought not only global devastation but also an unprecedented infodemic of false or misleading information that spread rapidly through online social networks. Network analysis plays a crucial role in the science of fact-checking by modeling and learning the risk of infodemics through statistical processes and computation on mega-sized graphs. This paper proposes MEGA, *M*achine Learning-*E*nhanced *G*raph *A*nalytics, a framework that combines feature engineering and graph neural networks to enhance the efficiency of learning performance involving massive graphs. Infodemic risk analysis is a unique application of the MEGA framework, which involves detecting spambots by counting triangle motifs and identifying influential spreaders by computing the distance centrality. The MEGA framework is evaluated using the COVID-19 pandemic Twitter dataset, demonstrating superior computational efficiency and classification accuracy.

## 1 Introduction

THE rapid global spread of the coronavirus disease 2019 (COVID-19) spawned a new crisis known as the COVID-19 infodemic, which the World Health Organization (WHO) declared equally devastating for COVID-19 pandemic control [1], [2], [3]. An infodemic refers to the overwhelming amount of false or misleading information that circulates in both digital and physical environments during a pandemic. In the early stages of a disease outbreak, when little is known about the disease, misinformation can be particularly damaging, leading to confusion and harmful outcomes. As shown in [4], the prevalence of false information on social media, which has a greater reach and impact than accurate information, highlights the critical need to address the infodemic problem urgently.

Fighting infodemics requires detecting spambots or unreliable sources that spread fake news or information with low credibility. According to [5], for every 21 tweets on Twitter, one is spam, and about 15% of active accounts are social bots. Managing infodemics also involves some form of source attribution, such as fact-checking, to root out malicious misinformation [1]. This can be accomplished by scoring users in an online social network with a reputation mechanism. For instance, the authors in [6] introduced the *Infodemic Risk Index (IRI)* to evaluate the likelihood of an infodemic occurrence in various nations. However, it is well-known that popular online social networks like Twitter have a high percentage of spambots. For example, it was alluded that over 10% of the 450 million monthly active Twitter users in 2022 were bots during Elon Musk’s takeover of Twitter. As viral messages in an infodemic can be caused by spambots (potentially spreading disinformation), assessing infodemic risks will require a dynamic approach to evaluate the credibility of a message (whether due to human or bot) and determine its reach.

Metrics that quantify infodemic risk at the individual user level can significantly enhance the science of fact-checking. Given its dependence on accurate and reliable information sources, fact-checking is a time-consuming process that involves validating or cross-checking claims from multiple sources. Proactive risk assessment can identify credible sources of news with sufficiently high scores and authenticate information in the form of a risk matrix for social listening. This allows infodemic risk managers to first scale up fact-checking and then mitigate the spread of misinformation or disinformation over the online social network.

In this paper, we propose a novel framework called MEGA, *M*achine Learning-*E*nhanced *G*raph *A*nalytics, for automated machine learning with mega-sized graph datasets. A novelty of MEGA is its use of automated feature-based vertex embeddings in graph neural networks (GNNs) to process massive graph datasets. MEGA comprises two major steps: *feature engineering* and *supervised learning*. MEGA first efficiently computes the most relevant features via automated feature engineering and then applies GNNs for supervised learning in downstream tasks. As an application, we show how to compute accurate infodemic risk scores using the COVID-19 pandemic Twitter dataset. The contributions of the paper are as follows:

- Our MEGA framework leverages feature-based vertex embeddings for GNNs to preserve important feature information for learning performance optimization.
- For breadth-first search (BFS) graph decomposition, we incorporate top-*k* ranking to reduce the number of BFS executions, significantly improving the feature engineering step in MEGA. This method, inspired in part by Tarjan’s BFS graph decomposition technique [7], [8], enhances the computation of network centrality-based problems involving massive graphs [9].
- We demonstrate MEGA’s superior performance over existing techniques by conducting extensive evaluations on two statistical graph problems (triangle motif counting and distance center computation) in the feature engineering step and two classification tasks (spambot detection and influential spreader identification) in the supervised learning step. The evaluations use a Twitter dataset containing over 1 million tweets related to the COVID-19 pandemic, collected over five months.

This paper is organized as follows. In Section 2, we review the related work in studies of infodemic risk management on social media. In Section 3, we present the MEGA framework and outline the triangle motif counting for spambot detection and distance center computation for influential spreader identification problems. In Sections 4 and 5, we demonstrate how MEGA solves the problems of triangle motif counting and distance center computation, respectively. In Section 6, we propose two feature-based vertex embedding methods with GNNs for spambot detection and influential spreader identification. We demonstrate the performance evaluation results and applications on COVID-19 infodemic risk management in Section 7. We conclude the paper in Section 8.

## 2 Related Work

Our work is related to automated machine learning (AutoML) for public healthcare applications [10]. AutoML is particularly useful for detecting spambots and influential spreaders due to its capability to automate the feature engineering process, identifying and extracting relevant features from massive graph datasets obtained from online social networks. The traditional approach of manual feature engineering has become increasingly impractical and time-consuming, especially in light of the rampant spread of misinformation. Automated feature engineering can also help to identify patterns in social media data that are not immediately obvious to human analysts. For instance, these techniques can help determine who is spreading pandemic-related misinformation to whom and how it is being disseminated across the network.

By automating the feature engineering of massive graphs, AutoML can reduce the risk of bias in data analysis and decrease the likelihood of overlooking key features that may not be apparent to human analysts [11], [12]. To detect spambots on social media, the work in [13] utilized unsupervised feature extraction and a clustering algorithm to distinguish between humans and bots. In [14], a BERT model was proposed to detect social bots whose activities are correlated with COVID-19-related fake news. The authors in [15] used unsupervised learning to identify graph features related to suspicious activity patterns for bot detection. The work in [16], [17] studied extracting graph features like vertex degree and triangle count to detect spam accounts.

Another feature engineering aspect of graph data is the influence of individual vertices in the graph. The work in [18] applied diffusion algorithms to quantify the influence of Twitter users using the Twitter follower graph. In [19], *K*-truss decomposition was used to locate influential vertices based on counting triangles. Other related works used network centrality to measure the influence of vertices [20], [21], [22], [23]. The work in [9] proposed the rumor centrality as the maximum-likelihood estimate of influential spreaders [24], which is the optimal solution when the given network is a tree. Epidemic centrality in [25] generalized the rumor centrality to graphs with cycles and can be computed by a message-passing algorithm. In contrast to finding the influential spreaders, the work in [23] considered the problem of preventing networks from cascading failures by protecting some vertices in advance.

To the best of our knowledge, our paper is the first to propose AutoML to jointly learn how to detect spambots and influential network users in order to compute infodemic risk scores in the context of the COVID-19 pandemic. This approach is inspired by the related work in [6] that analyzed COVID-19 pandemic-related Twitter data to compute an infodemic risk index for quantifying the exposure rate of aggregated Twitter users to unreliable news, but neither spambots nor influential users detection was considered in [6].

## 3 Machine Learning-Enhanced Graph analytics

In this section, a new MEGA framework is proposed and the triangle motif counting and distance center computation problems for infodemic risk management are formulated.

### 3.1 The MEGA Framework

Let *G* = (*V* (*G*), *E*(*G*)) be a simple undirected graph with vertex set *V* (*G*) and edge set *E*(*G*). The MEGA framework has two main stages:

#### Stage 1. Feature Engineering

##### 1) Graph Pruning

Decompose the given graph *G* into connected components to obtain the resultant graph *G*^*′*^. Set a threshold parameter *θ* and iteratively remove small-degree vertices from *G* until

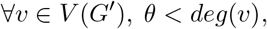

where *deg*(*v*) is the degree of vertex *v*. Vertex degree is used as a criterion in this step as it can be efficiently obtained without complex pre-computation compared to other vertex properties.

##### 2) Hierarchical Clustering

Cluster the vertices of a connected component hierarchically using the breadth-first search algorithm (BFS) [7], [8].

##### 3) Computing

Solve a network computational task based on the hierarchy property.

#### Stage 2. Supervised Learning

Apply a GNN with the feature-based vertex embedding for classification.

In this study, we consider undirected graphs over directed ones. This choice reduces complexity while preserving properties essential for efficient, accurate, and satisfactory analysis. The mutual relationships in undirected retweet networks allow for a more interpretable examination of spambot detection and influential user identification on Twitter. Conversely, using directed graphs would require adapting the methods to account for edge directionality, increasing complexity and complicating the interpretation of results. Fig. 1 presents an overview of MEGA, assessing the infodemic risk through spambot detection and influential spreader identification.

**Fig. 1.**
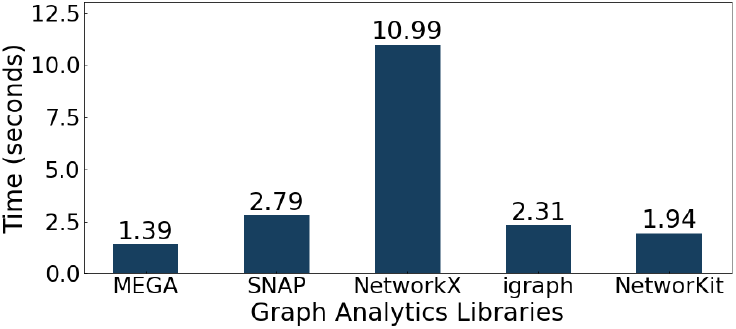
An overview depicting the interdependence among the tasks solved by the MEGA framework for infodemic risk management.

### 3.2 Motivation and Summary

To motivate our approach for solving the triangle motif counting and distance center computation problems for infodemic risk management, we present two examples to show the measures we compute can help in spambot detection and influential spreader identification for infodemic risk evaluation.

The work in [6] developed a risk index to quantify the exposure rate of a Twitter user to unreliable tweets shared by a specific class of users (partial IRI, (1)) or by any class of users (IRI, (2)):

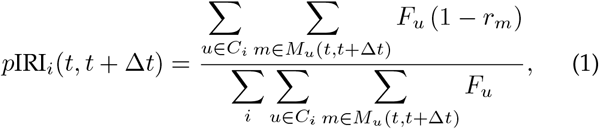

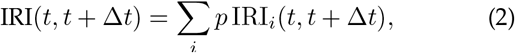

where *F*_*u*_ is the follower count of user *u*, defining the exposure of message *m* posted by *u* at time *t* in terms of potential visualizations by the first-order approximation, *r*_*m*_ is the reliability of *m* with a value of either 0 or 1, *M*_*u*_(*t, t* + ∆*t*) is the set of messages posted by *u* within a time window of length ∆*t*, and *C*_*i*_ (*i* = *V, U*) signifies user classes: verified (*V*) and unverified (*U*) accounts, as determined by the social platform. To evaluate the risk index from another perspective, we use spambot detection to assess *r*_*m*_ and influential spreader identification to measure *F*_*u*_ (see details in Section 7.5).

In our Twitter dataset of the COVID-19 pandemic (detailed in Section 7.1), we identify two categories of accounts: legitimate users and spambots. As seen in Fig. 5 (Section 7.3.3) and consistent with [16], legitimate users exhibit a higher triangle count than spambots. This observation leads to two conjectures: (a) the triangle distribution of legitimate users differs from that of spambots on Twitter, making the triangle motif count a robust spambot detection feature. Additionally, detection with content-based features can easily be evaded by filtering spam words, while the graph-based triangle count feature models real-world social relationships, making it harder to bypass. (b) Legitimate users tend to follow and retweet those with shared interests or similar views, leading to relationship formation and triangle creation in their networks.

Finding influential spreaders can help us maximize a tweet’s reach on Twitter based on past interactions (e.g., retweets). The motivation is similar to analyzing the effect of viral spreading and influence maximization [26], [27]. The goal is to find a small group of Twitter users (root vertices in a graph) to efficiently spread reliable tweets and maximize their reach over time. In [9], this was formulated as a maximum likelihood estimation problem (see Section 3.4), and rumor centrality was introduced to solve it optimally in *O*(*N*) time for infinite-size, degree-regular tree graphs with *N* vertices. However, the problem becomes computationally hard for non-tree graphs. A BFS heuristic algorithm for general graphs was proposed in [9], but it required *O*(*N* ^3^) time complexity due to BFS tree traversal starting from every vertex. Distance centrality is a proven suboptimal heuristic to approximate the optimal solution [9]. It achieves the optimal solution for degree-regular trees, like rumor centrality. Our MEGA framework computes the distance center faster by reducing the number of BFSs invoked on the graph, demonstrating that distance centrality is a more efficient measure of influence on Twitter.

### 3.3 Triangle Motif Counting for Spambot Detection

Triangle motif enumeration in spambot detection involves identifying and counting the triangle motifs.

#### Problem 1

(Counting triangle motifs). Given a simple undirected graph *G* = (*V* (*G*), *E*(*G*)), if there exist three vertices *v*_*i*_, *v*_*j*_ and *v*_*k*_ such that (*v*_*i*_, *v*_*j*_), (*v*_*i*_, *v*_*k*_) and (*v*_*j*_, *v*_*k*_) are all in *E*(*G*), then we say *v*_*i*_, *v*_*j*_, *v*_*k*_ form a triangle motif. The goal is to compute triangle count for each *v* ∈ *V* (*G*) by:

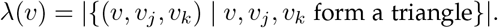

### 3.4 Distance Center Computation for Influential Spreader Identification

Identifying influential spreaders in online social networks is one of the most critical issues in statistical inference and infodemic risk management. This paper aims to find the *distance center* of a mega-sized graph accurately and efficiently.

#### Problem 2

(Computing the message source estimator). Assuming a tweet spreads on Twitter following the *susceptible-infected* (SI) model, how to accurately find the spreader in a spread graph where all vertices know the tweet’s origin?

The tweet spreading follows the SI model, in which once a vertex is “infected”, it stays in this state forever (i.e., once a tweet is retweeted, a relationship is built between the spreader and the retweeter, and this cannot be reversed). Let *V*_*I*_ denote the set of vertices such that each vertex in *V*_*I*_ has at least one infected neighbor. In each time slot, one vertex is uniformly chosen from *V*_*I*_ to be the next infected vertex. Given a snapshot of *N* infected vertices *G*_*N*_, we want to find

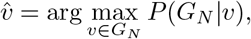

where 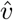 is the spreader and *P* (*G*_*N*_ |*v*) is the probability of having *G*_*N*_ under the SI model assuming *v* is the original message source. Note that we only focus on using the network topology of *G*_*N*_ to compute 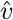 in this paper.

Let *G* = (*V* (*G*), *E*(*G*)) be a simple connected graph. Denote the shortest distance between two vertices, say *u* and *v*, by *dist*(*u, v*). The distance centrality of a vertex *v* ∈*V* (*G*), *S*(*v, G*), is defined as

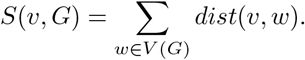

The distance center, *C*_*dist*_(*G*), of *G* is a vertex that has the smallest distance centrality.

## 4 MEGA for Triangle Motif Counting

In this section, we describe how MEGA solves the triangle counting problem for feature engineering (see Problem 1).

### 4.1 Graph Pruning

We remove small-degree vertices from the given graph *G* such that *θ < deg*(*u*) *< N* ^*′*^ − 1 for all *u* ∈ *V* (*G*^*′*^), where *G*^*′*^ is the pruned graph and *N′* is the cardinality of *V* (*G*^*′*^). If a vertex *u* is removed from *G*, then for each pair of vertices *a* and *b* both belonging to *N*_*G*_(*u*) = {*v* | (*u, v*) ∈ *E*(*G*)}, we check if (*a, b*) ∈ *E*(*G*). If (*a, b*) ∈ *E*(*G*), we count {*u, a, b*} as a triangle motif. Hence, there are 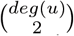 pairs need to be checked for each removed vertex *u*. If *N* ^*′*^ *<* 3, then MEGA ends here. Otherwise, we go to the next step.

In general, the threshold *θ* is a tunable parameter and its optimal value depends on the graph topology and the problem we wish to solve. We say *θ* is optimal if it can largely decompose a given graph and *θ* ≪*N* (see Appendix C). We provide bounds on the optimal threshold *θ*^∗^ for some special cases based on their inherent structures.

#### Lemma 1.

For any graph *G* = (*V* (*G*), *E*(*G*)), if all vertices with degree less than or equal to *θ* are removed to obtain the pruned graph *G*^*′*^ = (*V* (*G*^*′*^), *E*(*G*^*′*^)), then

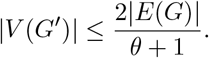

#### Theorem 1.

For any Barabási-Albert (BA) network *G* with parameters *m*_0_ and *m*, where *m*_0_ is the size of the initial complete network and *m* is the degree of each newly added vertex, the optimal threshold *θ*^∗^ to efficiently decompose *G* is *m*, and it can be computed by

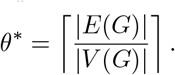

#### Lemma 2.

For any Erdős-Rényi (ER) random network *G*(*N, p*), by setting *θ*^∗^ = *k deg*_*avg*_, where *k* is a positive integer and *deg*_*avg*_ is the average degree of *G*, the size of the pruned graph decreases exponentially with increasing *k*.

#### Lemma 3.

For any planar graph *G*, the optimal threshold *θ*^∗^ to completely decompose *G* is 5.

### 4.2 Hierarchical Clustering

We leverage a hierarchical clustering algorithm proposed in [7], [8] to split vertices based on the BFS tree traversal. Note that *G*^*′*^ may not be a connected graph even if *G* was. Denote the *i*th connected component in *G*^*′*^ as *G*_*i*_. Then, 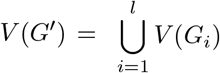, where *l* is the number of connected components (e.g., *l* = 1 if *G*^*′*^ is a connected graph). Let 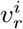 be the degree center (a vertex with maximum degree) of each connected component *G*_*i*_ for *i* = 1, …, *l*. Then, we use 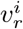 as the root of the BFS tree traversal for *G*_*i*_. For each *G*_*i*_, vertices are in the same cluster if their distances to 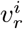 are identical. Hence, the *j*th cluster of *G*_*i*_, *K*_*i,j*_, is defined as

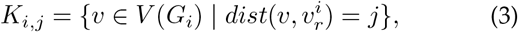

where 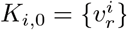 and 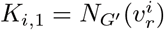.

### 4.3 Computing the Number of Triangle Motifs

Based on the BFS properties, for each motif {*v*_*i*_, *v*_*j*_, *v*_*k*_} in *G*^*′*^, there are only two possible structures:

### Inter-cluster motif

We call *v*_*i*_, *v*_*j*_, *v*_*k*_ an inter-cluster motif, if *v*_*i*_, *v*_*j*_ and *v*_*k*_ scatter in two neighboring clusters. In this case, one of the three vertices may be the root.

### Intra-cluster motif

If *v*_*i*_, *v*_*j*_ and *v*_*k*_ are in the same cluster, then we call {*v*_*i*_, *v*_*j*_, *v*_*k*_} an intra-cluster motif.

In the following, we describe how MEGA counts different structures of triangle motifs in the computing step.

#### 1) Rooted inter-cluster motifs

Similar to the graph pruning step, for each pair of vertices *a* and *b* both belonging to 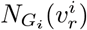, we check if (*a, b*) ∈ *E*(*G*_*i*_). Let 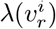 be the number of triangles incident to 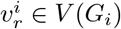. Then we have

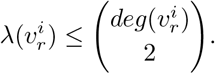

#### 2) Non-root inter-cluster motifs

For each vertex *v ∈ K*_*i,j*_, its upper neighbors, 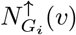, are difined by

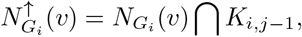

and its lower neighbors, 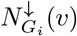, are difined by

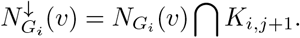

Thus, for each 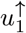 and 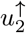 in 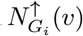, if 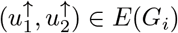, then 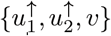 forms a motif. Similarly, for each 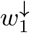 and 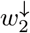 in 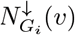, if 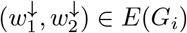, then 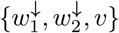 forms a motif. Let *λ*(*v*) be the number of triangles incident to *v ∈ V* (*G*_*i*_). Then we have

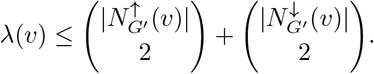

#### 3) Intra-cluster motifs

Let *G*_*i,j*_ = (*V* (*K*_*i,j*_), *E*(*K*_*i,j*_)) denote the induced subgraph of *G*^*′*^. Then, all intra-cluster motifs in *G*_*i,j*_ can be counted in a recursive manner (i.e., applying the feature engineering step of MEGA to count the number of triangle motifs in *G*_*i,j*_).

The triangle count of every vertex in *G* can be computed via steps 1-3 of the feature engineering phase, which is implemented by Algorithm 1 (see Appendix B).

## 5 MEGA for Distance Center Computation

In this section, we apply MEGA to compute the distance-based message source estimator (see Problem 2). In summary, the framework can find the distance center for tree graphs through the graph pruning step (Section V-A) alone. However, for cyclic graphs, the pruning step removes leaf vertices, after which the pruned graph undergoes further processing in the remaining two steps (Sections V-B and V-C) to find the distance center hierarchically using the BFS algorithm. Note that we only consider connected graphs in this section since we are ranking vertices in the same connected component to investigate message spreading.

### 5.1 Graph Pruning

We first remove the trivial vertices that have less chance to be the distance center in this step. Based on the statistical property of the data, we set the threshold *θ* to 1 so that all vertices in the pruned graph *G*^*′*^ have degrees larger than 1. Note that if we set *θ >* 1, *G*^*′*^ will become fragmented, which increases the computational complexity significantly.

Each vertex contains two parameters, subtree size *T* and sum of removed distances *D*. We use a rewriting system to update these two parameters for every vertex in the input graph *G*. Note that vertices receiving messages from a given vertex are referred to as parents *p*, and vertices for which a given vertex is parent are the children of that vertex, child(*p*). Initially, we set the subtree size to 1 and the sum of removed distances to 0 for every vertex. When a vertex *v* with degree 1 is removed from *G*, it sends the message of (*T* (*v*), *D*(*v*)) to its parent. Subsequently, the rewriting system updates the subtree size of each parent by

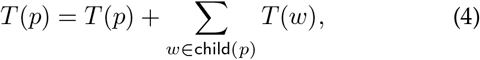

and the sum of removed distances by

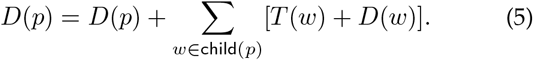

This step continues until all vertices in *G*^*′*^ have a degree greater than 1. Recall that the rewriting system in the *θ* = 1 graph pruning is equivalent to that in the *Election Algorithm* [28], and it can find the distance center in a tree within linear time complexity. However, social networks such as Twitter have more complex structures, where billions of users (vertices) can form millions of communities (cycles). Thus, finding the distance center in a tree is just a special case.

Let *G*^*′*^ = (*V* (*G*^*′*^), *E*(*G*^*′*^)) be the pruned graph with *N* ^*′*^ vertices. Then, *G*^*′*^ is a connected graph containing cycle(s) and *deg*(*v*) *>* 1 for all *v* ∈ *V* (*G*^*′*^). If *N* ^*′*^ *>* 1, we go to the next step. Otherwise, *G*^*′*^ is the distance center.

### 5.2 Hierarchical Clustering

In this step, we partition vertices in *G*^*′*^ into different clusters based on the BFS tree traversal. Let *v*_*r*_ be the root of the BFS for *G*^*′*^. Similar to (3), we use the distance between *v*_*r*_ and all other vertices in *G*^*′*^ to define each cluster. Since *G*^*′*^ must be a connected graph, we use *K*_*i*_ to denote the *i*th cluster in *G*^*′*^. We rewrite (3) as

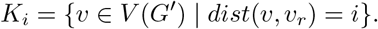

That is, a vertex *v* is in cluster *K*_*i*_ if its distance from *v*_*r*_ is *i*. We can then find the distance centrality of *v*_*r*_, *S*(*v*_*r*_, *G*^*′*^), by

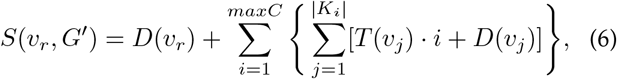

where *maxC* is the number of clusters in *G*^*′*^ and |*K*_*i*_| is the number of vertices in the *i*th cluster. Then, the total subtree size and the sum of removed distances of each cluster is 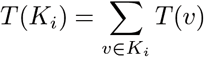 and 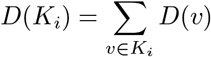, respectively.

The initial root *v*_*r*_ impacts not only the number of clusters, but also the lower bound computation of every vertex in the next step, affecting the overall performance of the feature engineering. Having more clusters results in greater differences between the lower bounds of vertices.We first identify a vertex *v* with the minimum degree in *G*^*′*^ to minimize the number of vertices in *K*_1_. Then, we choose the root *v*_*r*_ as a vertex with the maximum eccentricity, i.e., the farthest from *v*.

### 5.3 Computing the Distance-based Estimator

After selecting the root, we use the idea of computing top-*k* closeness centrality [29] as a basis, which is to trace the lower bound on the distance centrality of each vertex to avoid running BFS starting from every vertex. We denote the lower bound on the distance centrality of vertex *v* in *G*^*′*^ as *S*_*LR*_(*v, G*^*′*^). Note that if *S*(*v, G*^*′*^) *< S*_*LR*_(*u, G*^*′*^) or *S*(*v, G*^*′*^) ≤ *S*(*u, G*^*′*^) for all *u* ∈*V* (*G*^*′*^), then we can predict that *v* is the distance center of *G*^*′*^.

#### 5.3.1 Computing the Cluster-based Lower Bound

To calculate *S*_*LR*_(*u, G*^*′*^), where *u ∈ K*_*i*_, we establish Theorem 2 based on the triangle inequality to characterize the lower bound in terms of clusters, *T* (*K*_*i*_) and *D*(*K*_*i*_). Note that all information about the pruned vertices are already stored in *T* (*v*) and *D*(*v*) for all *v* ∈ *V* (*G*^*′*^).

##### Theorem 2.

Let *G*^*′*^ be a pruned graph and *u* ∈ *K*_*i*_. Then,

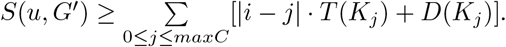

The two terms, *T* (*K*_*j*_) and *D*(*K*_*j*_), in Theorem 2 were computed in hierarchical clustering, and thus the complexity of computing the lower bound of vertex *u* is only a linear function of *maxC*. Note that in Theorem 2, the lower bound on the distance centrality of every vertex in the same cluster must be the same. Therefore, to tighten the lower bound of each vertex, we characterize two cases when |*i* − *j*| *<* 2:

*Case 1*: We have 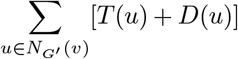 as the exact distance for *v* since *dist*(*u, v*) = 1 for all *u* ∈ *N*_*G′*_ (*v*).

*Case 2*: If *u* ∉*N*_*G′*_ (*v*), then we have 2 *T* (*u*) + *D*(*u*) as the lower bound for *v* since *dist*(*u, v*) must be at least 2.

#### 5.3.2 Computing the Distance Center

We propose Algorithm 2 to compute the distance center for the input graph (see Appendix B). We first compute *S*_*LR*_ for each vertex in *G*^*′*^ and insert it into a min-priority queue *Q*_*s*_ sorted in ascending order of *S*_*LR*_. Then, we use *DeQueue* to obtain the smallest *S*_*LR*_ in *Q*_*s*_ and check if *S*_*LR*_ = *S*. That is, if *S*_*LR*_(*v, G*^*′*^) is the smallest in *Q*_*s*_, then we do hierarchical clustering for *G*^*′*^ using *v* as the root and compute *S*(*v, G*^*′*^) with (6). If *S*(*v, G*^*′*^) = *S*_*LR*_(*v, G*^*′*^), then we conclude that *v* is the distance center of *G*^*′*^. Otherwise, we use *EnQueue* to put *S*(*v, G*^*′*^) into *Q*_*s*_ and repeat the whole process until we get the distance center of *G*^*′*^.

It is worth noting that if *v* is the root in the first hierarchical clustering and *S*(*v, G*^*′*^) is the smallest in *Q*_*s*_, then we only need to do BFS once. Otherwise, in the worst case, we have to do BFS for all vertices in *G*^*′*^. To further tighten the lower bound on the distance centrality for each vertex, we recompute the lower bound of every vertex in each BFS and update it if the bound is found to be tighter. That is, if the recomputed lower bound of a vertex appears to be larger than its current bound, then we update it. As we do more BFS, the lower bound should be updated to be tighter.

#### 5.3.3 Backtracking

Note that the distance center may be deleted in pruning for some graphs (e.g., pseudo-tree is a connected graph containing a single cycle). To resolve this issue, we establish Theorem 3 to characterize the cases when the distance center is in *G*^*′*^ and use Corollary 1 to find the distance center when it was deleted in pruning.

##### Theorem 3.

Let *C*_*dist*_(*G*) be the distance center of *G*. If *N* ^*′*^ ≥*N/*2, then *V* (*G*^*′*^) must contain *C*_*dist*_(*G*). If *N* ^*′*^ *< N/*2 and *C*_*dist*_(*G*) ∉ *V* (*G*^*′*^), then *C*_*dist*_(*G*) can be found in *O*(*dia*(*G*)) time, where *dia*(*G*) is the graph diameter of *G*.

##### Corollary 1.

If *N* ^*′*^ *< N/*2 and the unique vertex *v*^*′*^ with the minimum distance centrality in *V* (*G*^*′*^) is not *C*_*dist*_(*G*), then there is a unique path from *v*^*′*^ to *C*_*dist*_(*G*). Moreover, the distance centrality of each vertex along the path is monotonically decreasing.

Based on Corollary 1, MEGA is able to find *C*_*dist*_(*G*) in *O*(*dia*(*G*)) time after *C*_*dist*_(*G*) was deleted in graph pruning. In Appendix D, we demonstrate how MEGA efficiently finds the distance-based message source estimator for a general cyclic graph, without compromising on accuracy comparing with the BFS heuristic algorithm used in [9].

## 6 Supervised Learning WITH Graph Neural Networks FOR Infodemic Risk Management

In this section, we show how to leverage the deep learning architecture in graph to solve the spambot detection and influential spreader identification problems using the information obtained in the feature engineering step. In what follows, we first present two vertex embedding methods based on the number of triangles and distance centrality of each vertex respectively, and then show how to train the GNN models for classification in a supervised manner.

### 6.1 Vertex Embedding

Given a graph *G* = (*V* (*G*), *E*(*G*)), a traditional method for vertex embedding in GNN model is to use neural networks for vertices to aggregate information from their neighbors [30]. In the *k*-th layer of the GNN model, the update rules of a vertex *v* are given by

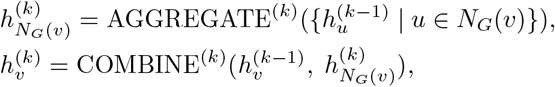

where *N*_*G*_(*v*) is the set of the neighbors of *v* ∈ *V(G)*, 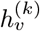 is the vertex features of *v* in the *k*-th layer or iteration, 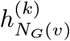 is an intermediate variable in the *k*-th layer or iteration, AGGREGATE(·) is a function (e.g., sum, mean, max-pooling and LSTM-pooling) to aggregate information from the neighbors of the vertex *v*, and COMBINE (*·*) is a function (e.g., summation and concatenation) to update the information of *v* combining the aggregated information from its neighbors and itself. The design of the two functions in GNNs is crucial and can lead to different kinds of GNNs.

However, the information aggregation step in the above embedding method treats each neighbor equally, and ignores the differences of important graph properties of these vertices. Meanwhile, all neighbors are sampled, which may lead to high computational complexity when the graph is huge. In fact, different vertices with different graph properties may transmit different amounts of information to their neighbors, so the influence of different neighbors of a vertex can be quite different depending on their graph properties. Instead of just exploiting the centrality information [31], we preserve the important feature information of data. In the following, we present two different feature-based vertex embedding methods to preserve the information of triangle count and distance centrality respectively.

#### Triangle-based Vertex Embedding

As we mentioned earlier, the number of triangles of a vertex is an important feature for us to judge whether a user account is a spambot. That is, a vertex with a larger number of triangles may convey more information for spambot detection. Thus, we propose considering the triangle count of the neighbors when aggregating the information. To do so, we sample the vertex’s neighbors with large triangle count, i.e., the top-*n* ranked vertices according to the triangle count out of all its neighbors. We also preserve the triangle count of each vertex during the aggregation, which is denoted by *t*_*u*_. Then, we construct the *k*-th layer to update a vertex *v* in the GNN model as

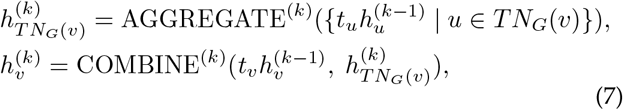

where *TN*_*G*_(*v*) is the set of the sampled neighbors of *v ∈ V* (*G*) based on the number of triangles of *v*’s neighbors. *Distance-based Vertex Embedding:* Computing the distance center is the first step to finding the message source, as in a real-world scenario, there is more than one spreader in a given network. Based on our simulation in Section 7.4.2, the average error between the real message sources and the top-*k* vertices with the smallest distance centrality is relatively small (within 3 hops) in the retweet network. Hence, we sample the vertex’s neighbors with small distance centrality when aggregating the information, i.e., the top-*n* ranked vertices with the smallest distance centrality out of all its neighbors. We also preserve the distance centrality of each vertex during the aggregation, which is denoted by *s*_*u*_. Then, we construct the *k*-th layer to update a vertex *v* in the GNN model as

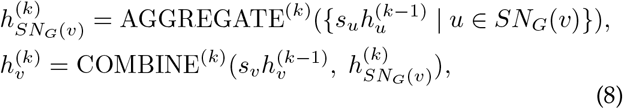

where *SN*_*G*_(*v*) is the set of the sampled neighbors of *v* ∈*V* (*G*) based on the distance centrality of *v*’s neighbors.

Fig. 2 shows the differences between traditional vertex embedding and our proposed triangle-based and distance-based vertex embeddings. To implement the feature-based vertex embeddings, we can simply adjust the adjacency matrix so that the original undirected graph becomes a directed one. We can then use the directed GraphSAGE [32] for the embedding.

**Fig. 2.**
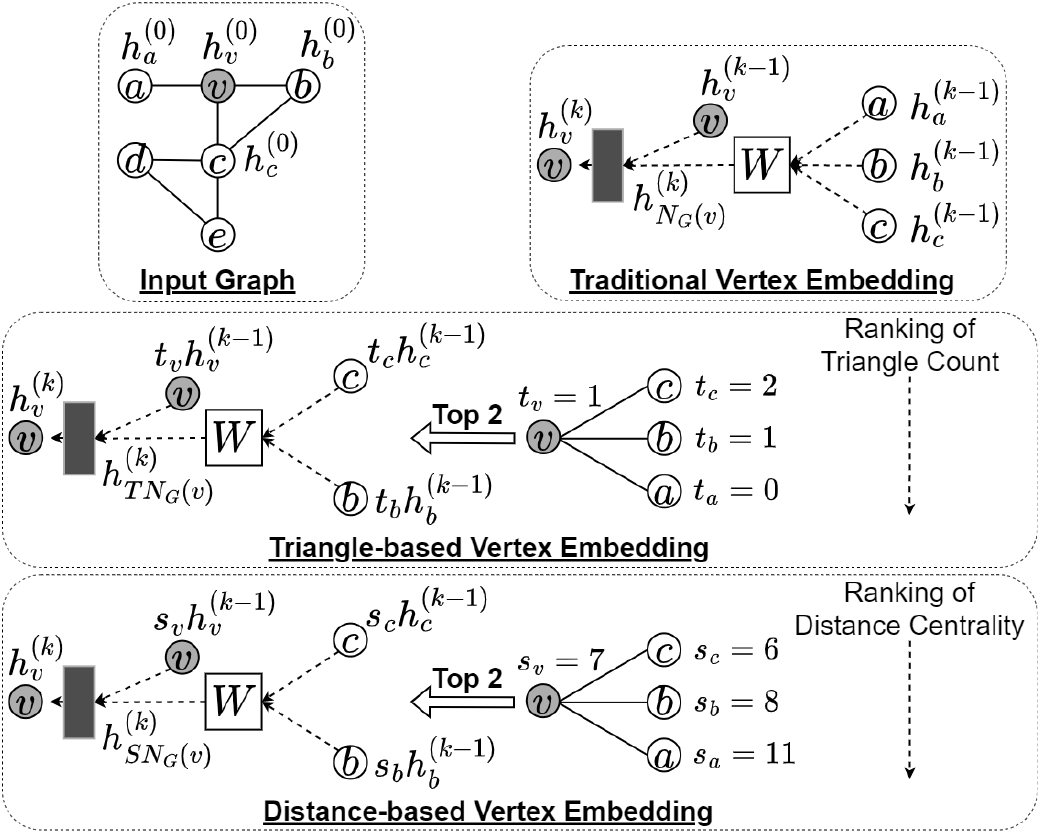
An example showing the differences between traditional vertex embedding and our proposed triangle-based and distance-based vertex embeddings, with vertex *v* (in gray) as the target for embedding. Gray rectangle boxes represent neural networks that combine aggregated information and information of *v*, while square boxes with a **W** matrix indicate trainable parameters for aggregating information from *v*’s neighbors. The traditional method aggregates information from all vertices, whereas our approach aggregates only the top 2 ranked neighbors.

### 6.2 Supervised Learning with Vertex Embedding

We propose a vertex-level classification model [33] that incorporates our vertex embedding methods. The model comprises two phases: message passing and classification. The message passing phase aims to learn a function generating vertex embeddings to capture vertex relationships based on the features of each vertex and its neighbors. The classification phase employs a neural network to classify each vertex by assigning a label. This approach can instantiate various models through different function settings for both phases.

For the message passing phase using the triangle-based vertex embedding, we elaborate the vertex embedding presented in (7) for each *v* in each layer *k* as follows:

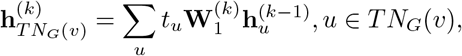

where 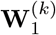 is a trainable parameter, and 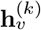 is the vertex features of *v* in the *k*-th layer. We set the initial feature 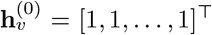. Then, the aggregated information and the information of *v* itself are combined as follows:

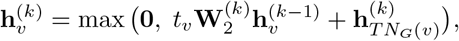

where 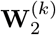 is a trainable parameter for the combination.

Similarly, for the message passing phase with the distance-based vertex embedding, the vertex embedding presented in (8) for each *v* in each layer *k* can be refined by

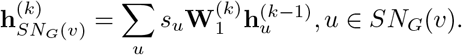

We also set the initial feature 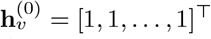. Then, the aggregated information and the information of *v* itself can be combined by

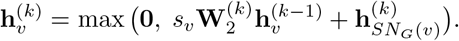

The classification phase remains the same for both vertex embedding methods. We predict the classification label 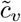 of *v* using a fully connected neural network, with the vertex feature as input:

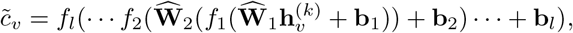

where 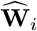 and **b**_*i*_ are trainable parameters, and *f*_*i*_ is the activate function, for *i* = 1, … *l*. To train the model, we select *Z* networks for the training set and use the cross-entropy loss function to update the learning parameters:

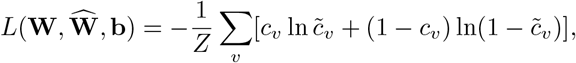

where *c*_*v*_ is the ground truth.

## 7 Numerical Simulations on COVID-19 Infodemic Risk Management

In this section, we first assess the performance of our MEGA framework on spambot detection and influential spreader identification on Twitter using the following criteria:

- The effectiveness of the feature engineering step, i.e., the speed of counting triangle motifs (Section 7.3.2) and the speedup of finding influential users (Section 7.4.1). We also provide more detailed experiments for the feature engineering step in Appendix E.
- The importance of the considered features, i.e., number of triangle motifs for spambot detection (Section 7.3.3) and distance centrality for influential spreader identification (Section 7.4.2).
- The performance of the GNN models in the supervised learning step for spambot detection (Section 7.3.4) and influential spreader identification (Section 7.4.3).

Then, we examine how spambot detection and influential spreader identification can be applied to assess the risk index for COVID-19 infodemic risk management (Section 7.5).

All the experiments are conducted on a 64-bit computer running a Windows 10 system with Intel(R) Core(TM) i7-9700K CPU 3.60GHz and 64.00 GB of RAM configuration.

### 7.1 Data Collection

On December 31, 2020, WHO issued its first emergency use validation for the Pfizer–BioNTech COVID-19 vaccine and emphasized the need for equitable global access. Since then, pro- and anti-vaccination comments, including inaccurate misinformation, have been circulating online. As such, our Twitter dataset of the COVID-19 pandemic was collected over a period of five months, covering the initial phase of the COVID-19 vaccine implementation. From January 1 to May 31, 2021, we scraped over 1 million tweets that contain any one of the following keywords: “COVID19”, “COVID”, “Coronavirus”, “Vaccine” and “Mask” (string matching is case-insensitive). We only search over English texts.

Due to the rate limit imposed by the Twitter API for data collection, we ignore tweets with fewer than 50 retweets. There are a total of 8,408 tweets recorded. For each tweet, we mark down the author and all the users who have retweeted the original tweet. We then build a (simple undirected) retweet network with 314,376 vertices and 519,178 edges. Each vertex represents a Twitter user, and the edge between two users means that they have retweeted each other at least once. The retweet network may not be a connected graph. The COVID-19 Twitter dataset is available at: https://github.com/MEGA-framework/Twitter-Dataset.

We consider the retweet network instead of other types of Twitter graphs as retweet data is more robust than the data of other social reactions (e.g., replying and following). The action of retweeting is often an approval or recognition of others’ thoughts, while following and replying have been shown to be of a different nature [34]. For example, spam contents are less likely to be retweeted by other users. Therefore, retweet data is more relevant in our study.

### 7.2 Baseline Models and Evaluation Metrics

To evaluate the classification performances of our proposed MEGA framework on spambot detection and influential spreader identification, we use the following three classification models as the baseline, namely Support Vector Machine (SVM), Logistic Regression (LR), and Random Forest (RF). We use two widely adopted evaluation metrics to assess the classification performance: accuracy *A* and F1-score *F* 1. We denote the true positive as *TP*, true negative as *TN*, false positive as *FP*, and false negative as *FN*. Then, we have:

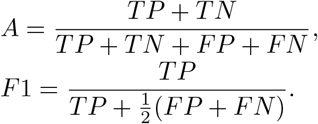

### 7.3 Results of Spambot Detection

In this section, we evaluate the performance of MEGA on spambot detection.

#### 7.3.1 Ground Truth Construction

The Twitter API does not provide any tools or information for us to determine if a user is a spambot, so we have to construct the ground truth of our self-collected Twitter dataset. We train our MEGA and the baseline models using the Twitter Social Honeypot Dataset [35], which has 22,223 spambots and 19,276 legitimate users. As this dataset is relatively old, we manually verify about 25% of the users. In the training process, we use 80% of the data as the training data and the remaining 20% as the test data with five-fold cross-validation. The performances are listed below:

|  | MEGA | SVM | LR | RF |
| --- | --- | --- | --- | --- |
| Accuracy | 94.3% | 82.3% | 86.9% | 87.6% |

We then apply the trained models to label our self-collected dataset based on the majority vote. For example, if SVM and LR both vote for “legitimate user” (0.823 + 0.869 = 1.692) and MEGA and RF vote for the opposite (0.943 + 0.876 = 1.819), then the label should be “spambot”. To further validate the results, we also manually check about 10% of users.

#### 7.3.2 Triangle Motif Counting

We compare the MEGA framework with the following four benchmarks based on the running time of each algorithm for triangle motif counting:

- Stanford Network Analysis Platform (SNAP) [36]: It enhances flexibility and efficiency in triangle motif counting by relying on the underlying graph data structure.
- NetworkX [37]: It uses the underlying graph data structure and vertex degree properties for triangle counting.
- igraph [38]: It first lists all triangles, then counts the number of triangles a vertex belongs to.
- NetworKit [39]: It uses a triangle listing algorithm from [40] to accelerate the triangle counting process.

These four benchmarks are single machine, single thread graph analytics libraries that could achieve competitive performance even compared to distributed graph frameworks.

In Fig. 3, we see that MEGA with optimal threshold *θ*^∗^ = 15 performs better than the above benchmarks on the retweet network for triangle motif counting. Thus, if we could find *θ*^∗^ that leverages the structure of the graph in advance, then MEGA can complete the counting task in graph pruning with *O*(*N*) time and beats other algorithms.

**Fig. 3.**
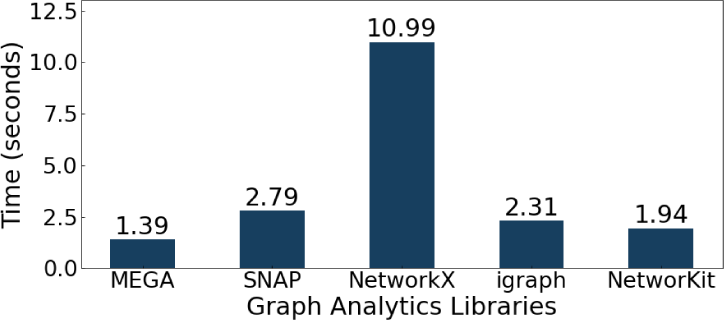
Comparison between MEGA and the benchmarks on triangle motif counting. The y-axis is the running time in seconds, and the x-axis is the graph analytics libraries we considered in the experiment. The number on top of each bar is the exact running time of the libraries.

The threshold *θ* is a critical parameter in graph pruning. Once *θ* is set, there are at most 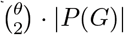 pairs of vertices need to be checked, where |*P* (*G*) | is the cardinality of the set of pruned vertices of *G*. Hence, a larger *θ* leads to a longer time spent for graph pruning. On the other hand, if more vertices are pruned off from *G*, then the graph becomes more fragmented. Thus, there is a trade-off between graph pruning and hierarchical clustering, and it depends on the threshold *θ*. Fig. 4 shows how *θ* affects the computation time of MEGA on the retweet network. We see that MEGA with the optimal threshold *θ*^∗^ = 15 performs significantly better.

**Fig. 4.**
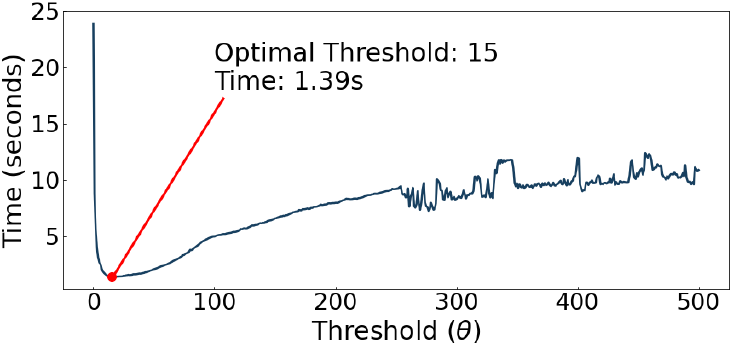
The performance of triangle motif counting with different thresholds *θ* ranging from 0 to 500. The y-axis is the running time in seconds, and the x-axis is the value of *θ*. The threshold *θ* = 15 gives the best performance with running time 1.39s.

**Fig. 5.**
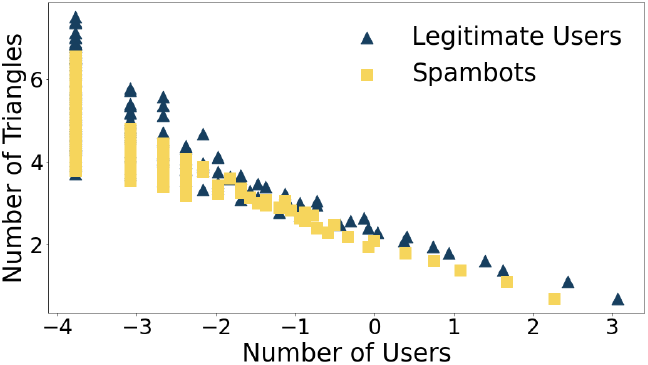
Comparison of the triangle count distributions between legitimate users and spambots. A log scale is used for both x and y axes.

#### 7.3.3 Feature Correlation

To verify that the triangle count is a robust and relevant feature for spambot detection in our Twitter dataset, we rank the importance of the following features:

##### Graph-based

(i) Triangle count, (ii) Eccentricity, (iii) Degree centrality, (iv) Betweenness centrality, (v) Distance centrality, (vi) PageRank centrality, (vii) Eigenvector centrality, (viii) Harmonic centrality, and (ix) Clustering coefficient.

##### User-based

(i) Number of followers *f*_*in*_, (ii) Number of friends (i.e., number of accounts a user is following) *f*_*out*_, and (iii) Reputation 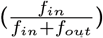.

##### Content-based

(i) Vectorized text of the original tweet, (ii) The length of the original text, (iii) Average word length of the original text, (iv) Vectorized text of the tweet without stop words (i.e., filtered text), (v) The length of the filtered text, (vi) Average word length of the filtered text, and (vii) The length ratio of filtered text to original text.

We compute the chi-squared statistics between each feature and the class label to rank the importance of the features for spambot detection. In Table 1, the triangle count is ranked as feature number 1 out of 19. Such a remarkably high correlation makes this feature well worth using for spambot detection. In Fig. 5, we also see that the triangle distribution of legitimate users is different from that of spambots.

**TABLE 1.** Top 10 important features for spambot detection.

| Rank | Feature |
| --- | --- |
| 1 | Triangle count |
| 2 | Reputation |
| 3 | Betweenness centrality |
| 4 | Degree centrality |
| 5 | Harmonic centrality |
| 6 | The length of the original text |
| 7 | The length of the filtered text |
| 8 | Eigenvector centrality |
| 9 | Average word length of the original text |
| 10 | Vectorized text of the original tweet |

**TABLE 2.**
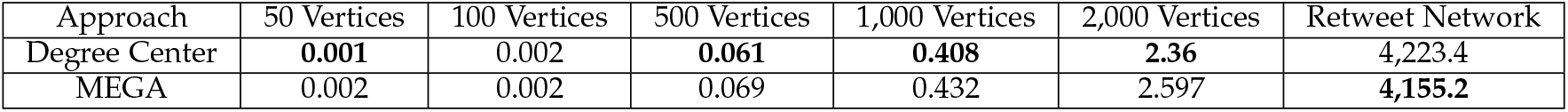
Comparison of the running time (in seconds) between MEGA and the small-world analysis approach using degree center as the root of the BFS in hierarchical clustering. The better-performing method is indicated in bold text.

| Approach | 50 Vertices | 100 Vertices | 500 Vertices | 1,000 Vertices | 2,000 Vertices | Retweet Network |
| --- | --- | --- | --- | --- | --- | --- |
| Degree Center | **0.001** | 0.002 | **0.061** | **0.408** | **2.36** | 4,223.4 |
| MEGA | 0.002 | 0.002 | 0.069 | 0.432 | 2.597 | **4,155.2** |

**TABLE 3.**
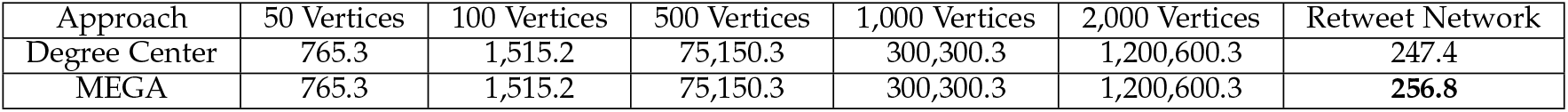
Comparison of the speedup between MEGA and the small-world analysis approach using degree center as the root of the BFS in hierarchical clustering. The better-performing method is indicated in bold text.

| Approach | 50 Vertices | 100 Vertices | 500 Vertices | 1,000 Vertices | 2,000 Vertices | Retweet Network |
| --- | --- | --- | --- | --- | --- | --- |
| Degree Center | 765.3 | 1,515.2 | 75,150.3 | 300,300.3 | 1,200,600.3 | 247.4 |
| MEGA | 765.3 | 1,515.2 | 75,150.3 | 300,300.3 | 1,200,600.3 | **256.8** |

**TABLE 4.** Comparison of the average error in identifying prominent vertices in small-world networks using distance centrality (MEGA), betweenness centrality, and degree centrality as the metrics. The best approach is indicated in bold.

| Approach | 50 Vertices | 100 Vertices | 500 Vertices | 1,000 Vertices | 2,000 Vertices |
| --- | --- | --- | --- | --- | --- |
| Distance Centrality (MEGA) | **1.29** | **2.11** | **2.37** | **2.27** | **2.59** |
| Betweenness Centrality | 1.31 | 2.21 | 2.44 | 2.32 | 2.63 |
| Degree Centrality | 1.32 | 2.18 | 2.47 | 2.36 | 2.60 |

**TABLE 5.** Comparison of the average running time (in seconds) in identifying prominent vertices in small-world networks using distance centrality (MEGA), betweenness centrality, and degree centrality as the metrics. The best approach is indicated in bold.

| Approach | 50 Vertices | 100 Vertices | 500 Vertices | 1,000 Vertices | 2,000 Vertices |
| --- | --- | --- | --- | --- | --- |
| Distance Centrality (MEGA) | 0.0021 | 0.0024 | 0.0028 | 0.0023 | 0.0027 |
| Betweenness Centrality | 0.0027 | 0.0031 | 0.0033 | 0.0029 | 0.0030 |
| Degree Centrality | **0.0012** | **0.0012** | **0.0013** | **0.0011** | **0.0012** |

#### 7.3.4 Classification Performance

We compare MEGA, which applies GNN with the triangle-based vertex embedding method, with the baseline models for spambot detection. In Fig. 6(a), we see that MEGA achieves the highest accuracy of 92.1% among all the considered learning models. Meanwhile, the F1-score of MEGA is also the highest (89.3%) in Fig. 6(b). This shows that our triangle-based vertex embedding is robust in spambot detection.

**Fig. 6.**
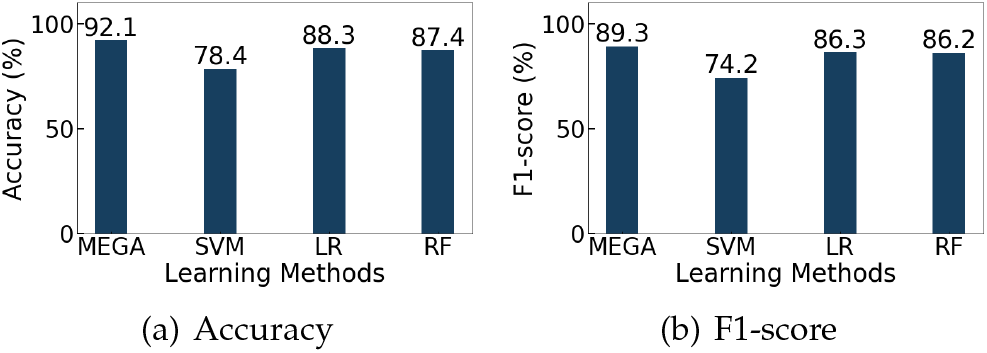
Classification performances of MEGA and the baseline models on spambot detection.

### 7.4 Results of Influential Spreader Identification

In this section, we assess the performance of MEGA on influential spreader identification.

#### 7.4.1 Distance Center Computation

We compare the MEGA framework with the algorithm presented in [29] for distance center computation. The work in [29] has been shown to outperform other existing algorithms for finding the exact and approximate top-*k* closeness centrality. In addition to the running time, we also assess the performance improvement based on the algorithmic speedup. The *speedup* suggested by [29] is given by 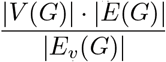, where |*E*_*v*_ (*G*)| is the number of edges that have been visited by an algorithm in *G*.

We extract the largest (strongly) connected component of the retweet network for evaluation. It has 308,147 vertices and 513,399 edges, and the graph diameter is 18. The solution in [29] uses the level-based and neighborhood-based lower bound (*NBBound*) to compute the top-*k* closeness centrality of the input graph *G* directly. It calculates the lower bound for every vertex in *G* that may also take those trivial vertices (leaves) into consideration when computing the top-*k* closeness centrality. If the number of vertices and edges we need to handle is always less than the NBBound after pruning, the number of edges visited by our algorithm must be less than the NBBound for every BFS. Thus, the more vertices we have removed in pruning, the larger the speedup we can reach. In this case, the running time must also be lesser for every BFS. In the retweet network, more than 73% of vertices were removed in pruning. Thus, in Fig. 7, we see that the running time and speedup of MEGA are better than the NBBound.

**Fig. 7.**
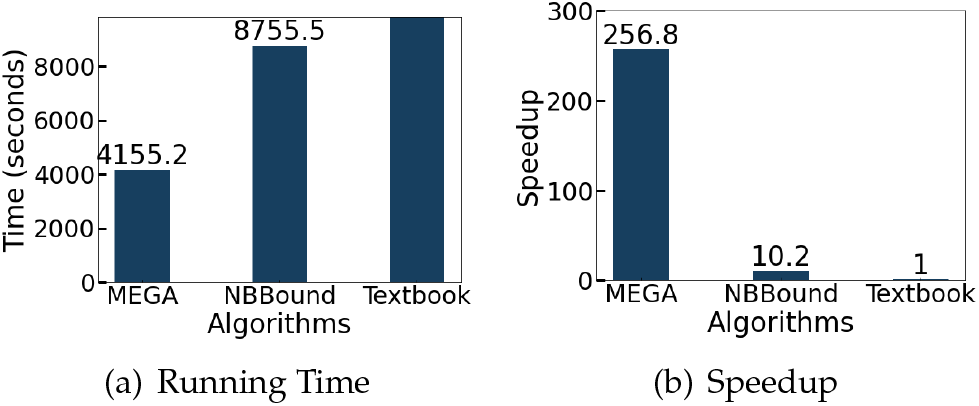
Comparison between MEGA and the top-*k* closeness centrality algorithm proposed by [29] on distance center computation based on the running time and speedup. We also show the performance of the textbook algorithm on this problem.

#### 7.4.2 Feature Correlation

We verify the hypothesis that the distance centrality is an essential feature for finding multiple message sources. Here, we also use the largest (strongly) connected component of the retweet network for evaluation. Since there are 5,195 tweets in this network, we identify top 5,195 vertices with smallest distance centrality and largest rumor centrality respectively. Fig. 8 shows the average error of using the distance-based feature is only 2.37 hops from the real source of 5,195 tweets, while the rumor centrality produces an error of 2.66 hops. Thus, the distance centrality is a more reliable feature in finding the message sources in Twitter than the rumor centrality.

**Fig. 8.**
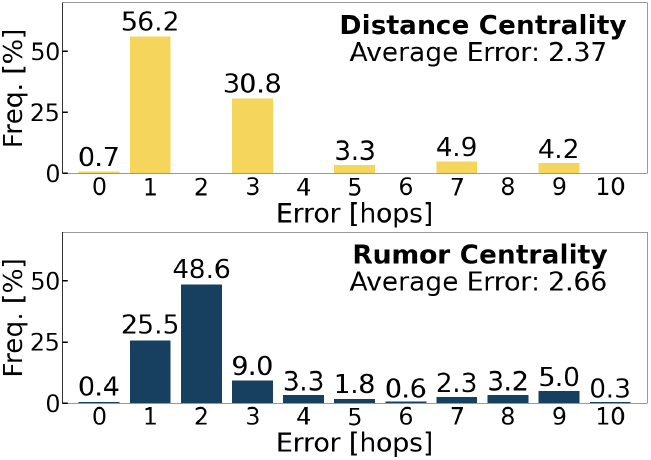
Histograms of the error for finding multiple message sources using the distance centrality and rumor centrality on the largest connected component of the retweet network.

#### 7.4.3 Classification Performance

we compared MEGA using GNN with distance-based vertex embedding to baseline models for identifying multiple message sources. Fig. 9 shows that the average performance of all the learning models is not as good as in spambot detection because the number of spreaders in the dataset is much lesser than those who retweet. Here, MEGA has the highest accuracy of 80.4% and highest F1-score of 77.2% among all the considered learning models, showing that our distance-based vertex embedding is another promising approach in influential spreader identification.

**Fig. 9.**
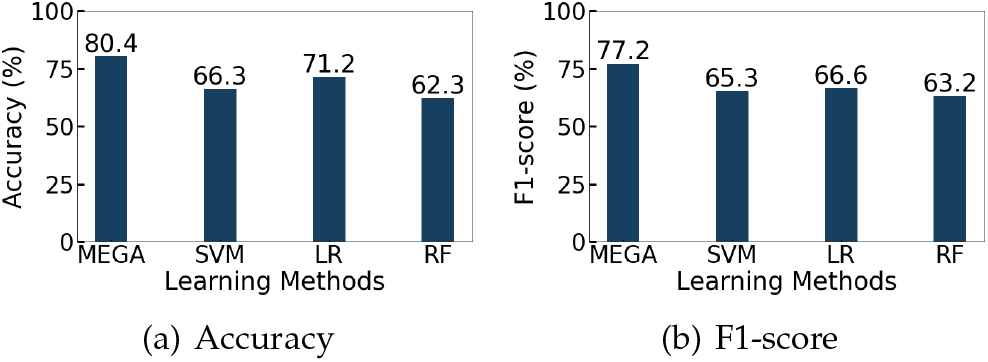
Classification performances of MEGA and the baseline models on influential spreader identification.

### 7.5 Infodemic Risk Index Evaluation

In this paper, we propose to solve the problems of spambot detection and influential spreader identification for evaluating the IRI. Recalling the IRI equations (1) and (2) in Section 3.2, we can determine the message reliability *r*_*m*_ of user *u* by identifying if *u* is a spambot such that

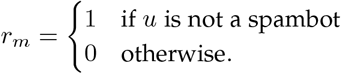

Let *S*_*m*_(*u*) be the distance centrality of a message *m* posted by *u* in the retweet network (i.e., the shortest distance for the message to travel from *u* to all other users in the retweet network). Then, we can redefine the exposure of a message posted by *u, K*_*u*_, by 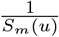. The idea is that, for a given graph, if a message can easily reach all other users (i.e., small distance centrality), then its exposure should be higher. The definition of the original IRI is more related to severity of the spread of rumors related to COVID-19. The fake news may be circulated and widely spread depending on the severity of the rumor spreading events. Thus, finding the sources of rumors is valuable to policymakers for infodemic control. We replace the unverified and verified accounts with spreaders and retweeters.

The original message reliability *r*_*m*_ is determined by identifying if *u* is an unreliable source spreading fake news. Therefore, to compute the original IRI for our analysis, we have to identify if the tweet content is real or fake. We use the Support Vector Machine (SVM) model with the training data containing 10, 700 fake and real news related to COVID-19 from [41] to identify whether a tweet is a piece of fake news (misinformation from unverified sources) or real news (useful information from reliable sources such as government authorities and news channels). The training data was collected from various fact-checking websites and social media, and the veracity of each news was manually verified in [41]. Using the class-wise balanced training data with the term frequency-inverse document frequency (TF-IDF) features, the SVM classifier has a test accuracy of 93.32% for fake news detection. We then apply the trained SVM classifier to our Twitter dataset. To further validate the classification results, we also use Google Fact Check Explorer and Coronavirus Rumor Control system to manually verify the veracity of each tweet. Personal comments about COVID-19 without a reliable source are considered fake news. There are totally 2,599 real news and 5,809 fake news. We give examples of real and fake news of our Twitter dataset below:

*Fake News*: *None of these masks will help you against #Covid 19 #COVID2019*.

*Real News*: *70 Covid-19 vaccines are under development, with 3 in human trials, the @WHO says #CoronavirusPandemic*.

Fig. 10 shows the modified IRI worldwide. We also analyze the original and modified IRI of top 15 countries with the most retweets, namely the United States (US), India (IN), United Kingdom (UK), Australia (AU), Canada (CA), Malaysia (MY), South Africa (SA), Pakistan (PAK), Ireland (IE), Philippines (PH), China (CN), Thailand (TH), Singapore (SG), Nigeria (NG), and France (FR). In Fig. 11 and 12, we see that although the original IRI is averagely larger than the modified IRI, but the distributions are quite the same. This result is as expected, as spambots usually have a large number of fake followers to increase their influence (i.e., larger *K*_*u*_ in the original IRI), but their messages indeed are unlikely to be retweeted by all the followers (i.e., lower *K*_*u*_ in the modified IRI). The original IRI is based on the number of fake news, while the modified IRI is based on the number of spambots. Therefore, having similar IRI distributions implies that most fake news comes from spambots.

**Fig. 10.**
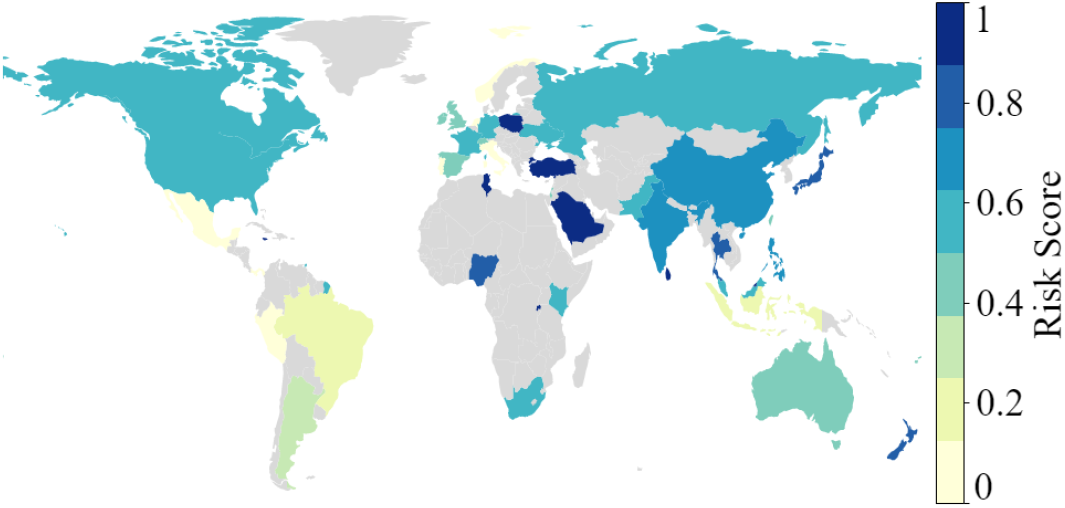
The modified IRI worldwide. The modified IRI of each country aggregated over five months, is color-coded on the map. The bar indicates different exposure levels; 1 means high exposure, and 0 means low exposure.

**Fig. 11.**
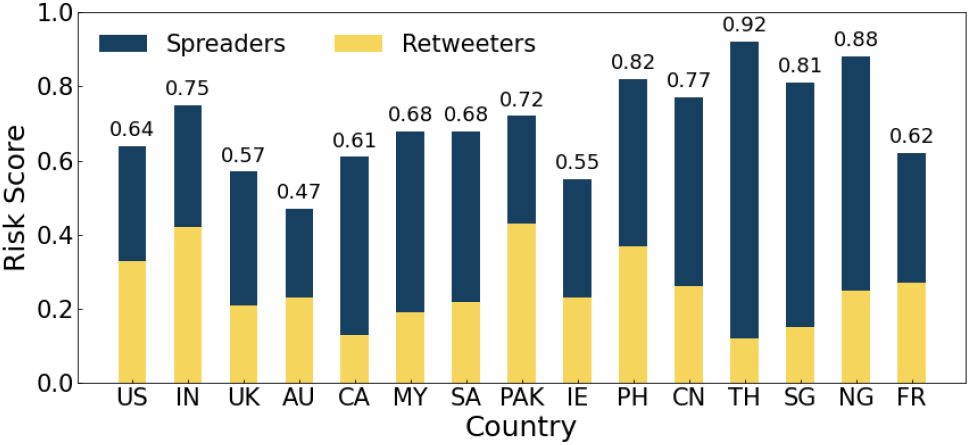
Original IRI of 15 considered countries. We compute the original IRI based on (1) and (2).

**Fig. 12.**
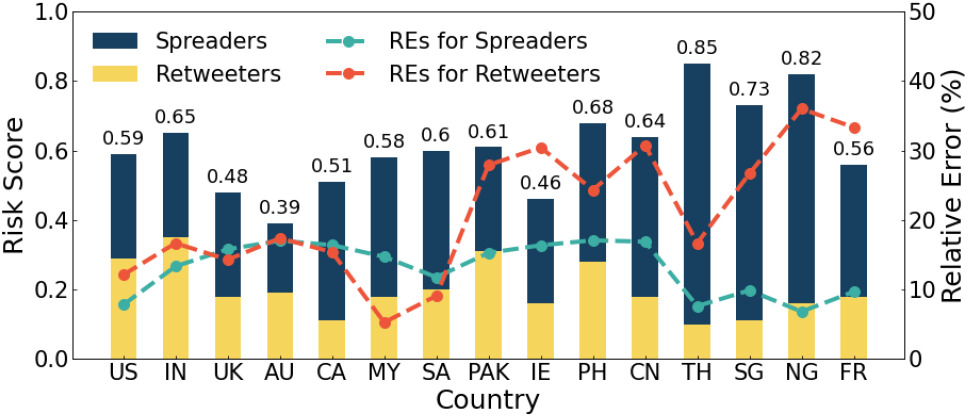
Modified IRI of 15 considered countries. We compute the modified IRI based on the changes in Section 7.5.

Regarding the COVID-19 vaccination status, the countries with high IRI values have low vaccination rates (see Fig. 13). During the initial phase of the COVID-19 vaccine implementation, there was a vast amount of unfounded rumors about deaths after receiving a particular COVID-19 vaccine or even after administering two injections of a particular vaccine, it cannot protect against certain virus variants. This situation is reasonable as the general public had little knowledge of the vaccine and the virus. More accurate information from experts and the WHO boosts confidence in COVID-19 vaccines, leading to higher vaccination rates in communities. However, the media environment is quite different across different countries. Fake news and spambots may not easily be verified due to the high cost of fact-checking. This explains why some countries, such as South Africa and Pakistan, with low online discourse, have a relatively large IRI and low vaccination rate.

**Fig. 13.**
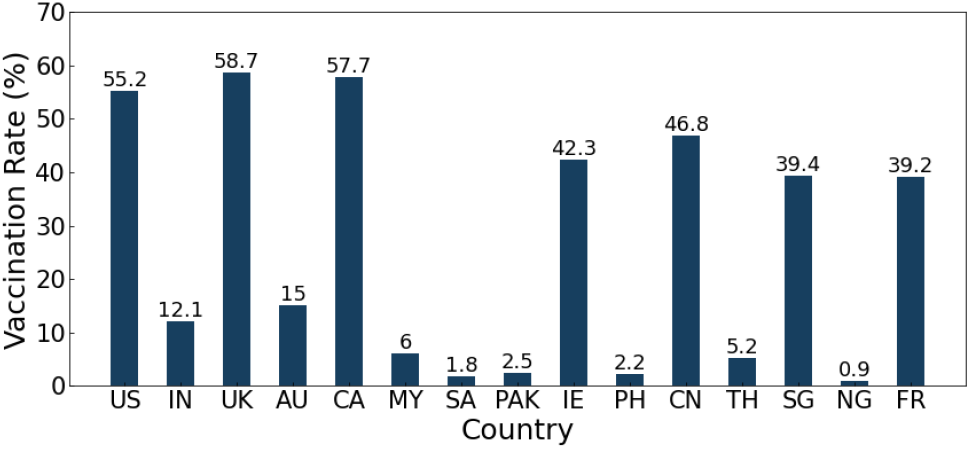
Vaccination rate of 15 considered countries. The vaccination rate is based on the percentage of the population that received at least one dose of a COVID-19 vaccine.

## 8 Conclusion

Our proposed system, MEGA (Machine learning-Enhanced Graph Analytics), employs graph neural networks to learn the underlying statistics of graph data and compute infodemic risk scores for users in online social networks. By combining graph analytics with AutoML to detect spambots and influential spreaders, MEGA outperforms existing techniques. Specifically, we demonstrate how optimizing hyperparameters in vertex embedding of graph neural networks preserves subgraph features and enables accurate computation of distance centrality. MEGA showcases how leveraging statistical features of graph datasets can facilitate efficient feature engineering, reduce the overall computational complexity of processing massive graphs, and compute accurate infodemic risk scores.

In our current work, we utilize triangle count and distance centrality for ranking the importance of vertices in vertex embedding. However, there is a need to preserve more than just one feature for embedding. In future research, we plan to extend the MEGA framework to address other computationally challenging graph representation learning problems, and improve the pipeline from social listening to infodemic risk measure computation for automated fact-checking. Incorporating advanced language models such as ChatGPT [42] into future research also offers a promising direction for enhancing the accuracy and efficiency of automated fact-checking and infodemic risk management.

## Data Availability

No

## Appendixes

This section contains the appendixes of the paper. In Appendix A, we provide the proofs of Lemma 1 – 4 and Theorem 1 – 2 of the main paper. We present the pseudocodes for the triangle motif counting algorithm and distance center computing algorithm of the feature engineering step of MEGA in Appendix B. In Appendix C, we analyze the time complexity of the proposed triangle motif counting algorithm of our MEGA framework. In Appendix D, we provide an illustrative example to demonstrate how the feature engineering step of MEGA computes the distance center for single message source detection. In Appendix E, we provide more detailed experiments to assess the performance of the feature engineering step of MEGA for triangle motif counting and distance center computation.

### APPENDIX A

#### Proofs

##### A.1 Proof of Lemma 1

*Proof*. According to the *degree sum formula*, the sum of the degrees of all vertices in a finite graph is twice the number of edges in that graph. Since the degree of every vertex in *G*^*′*^ must be at least *θ* +1, the sum of the degrees of all vertices in *G*^*′*^ must be greater than or equal to *θ* + 1 times the number of vertices in *G*^*′*^. That is,

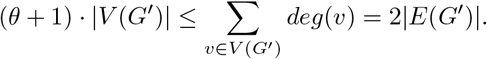

Since *G*^*′*^ is a pruned graph of *G*, the number of edges in *G*^*′*^ must be less than or equal to that in *G*. Hence, we have

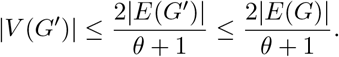

##### A.2 Proof of Theorem 1

*Proof*. We denote a BA network with parameters *m*_0_ and *m* as *G* and the initial complete network of *m*_0_ vertices in *G* as 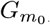, where *m*_0_ *> m*. Then, we have

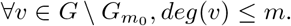

If we set *θ*^∗^ = *m*, it is obvious that the size of the pruned graph must be equal to *m*_0_ which is the size of 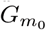. If *m* = *m*_0_ − 1, then we can completely decompose *G*. □

##### A.3 Proof of Lemma 2

*Proof*. Let *G*(*N, p*) be an ER random network. Then, the distribution of the vertex degrees is Poisson as the graph size *N* goes to infinity. Thus, we can use the Chernoff bound [43] to compute the upper bound of the size of the pruned graph. Let *v* be any vertex in *G*(*N, p*) and *θ*^∗^ = *kNp* be the optimal threshold for pruning, where *k* is a positive integer, then we have

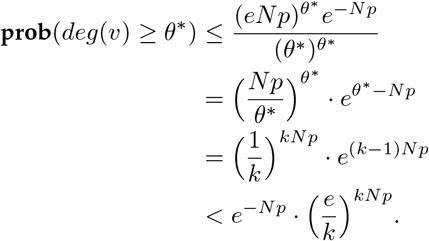

Therefore, we can have a simple upper bound for the size of the pruned graph, *N* ^*′*^, as follows:

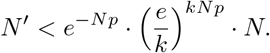

Note that for an ER random network *G*(*N, p*), the value *Np* is the average degree of the graph, which is a constant. Hence, the size of the pruned graph decreases exponentially as *k* increases. It is worth noting that the key property used to prove Lemma 2 is that the probability distribution of a vertex has degree *k* satisfies the Poisson distribution when the size of the graph is large enough. □

##### A.4 Proof of Lemma 3

*Proof*. Let *G* = (*V* (*G*), *E*(*G*)) be a planar graph. It is known that the average degree of *G* is strictly less than 6. It implies that there must be at least one vertex with degree less than or equal to 5 in *G*. After removing such vertices from *G*, there is always at least one new vertex to prune since any subset of a planar graph is also a planar graph. Ultimately, *G* can be completely decomposed. □

##### A.5 Proof of Theorem 2

*Proof*. Let *v* ∈ *K*_*j*_, where *j* is an integer such that 0 ≤ *j* ≤ *maxC*. Let 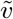 denote the set of pruned vertices such that *v* is the ancestor of these pruned vertices (e.g., 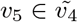 in Fig. 14). First, we consider the distance from vertex *u* to all the vertices in 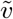 including *v* itself. Without loss of generality, we assume that the common ancestor *w* of *u* and *v* is in level *h*. Then, we have

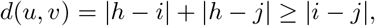

where the equality holds if *u* = *w* or *v* = *w*. Then, for other vertices in 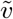, we have

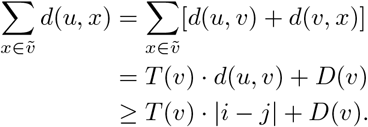

**Fig. 14.**
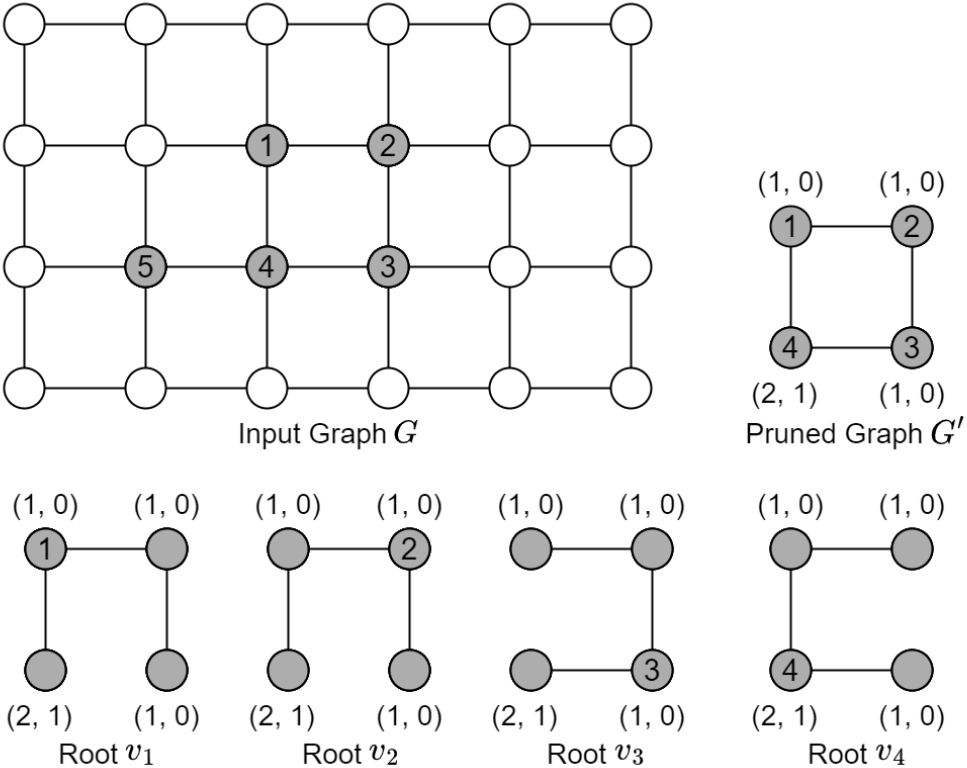
Example illustrating how MEGA finds the distance-based message source estimator of a spread graph. There are totally 5 vertices received the message (in gray color). After graph pruning, we have a pruned graph *G*^*′*^. We then select *v*_1_ to *v*_4_ as the root in hierarchical clustering respectively and see how many BFSs we have to build for each root.

Hence, we have

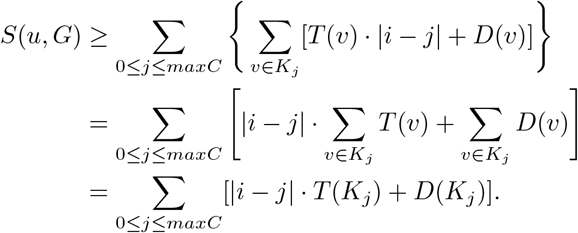

##### A.6 Proof of Theorem 3

*Proof*. First, we show that *G*^*′*^ must contain *C*_*dist*_(*G*) whenever *N* ^*′*^ *≥N/*2. We prove this part by contradiction. Assume that *N* ^*′*^ *≥N/*2 and the distance center *v*_*c*_ *∈G \G*^*′*^. To prove this theorem, we define an edge (*u, v*) as a bridge if the removal of (*u, v*) disconnects *G*. Assume (*u, v*) is a bridge. Let 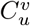 and 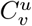 denote the connected component containing *u* and *v* respectively after removing (*u, v*).

###### Lemma 4.

If edge (*u, v*) is a bridge in *G*, then we have

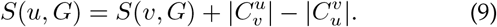

*Proof*. Let *u* and *v* be two vertices of *G* such that (*u, v*) is a bridge in *G*. Then, for any vertex 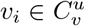 and 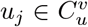, we have

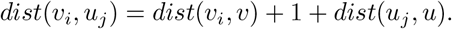

Hence, the distance centrality of *u* in *G, S*(*u, G*), can be rewritten in terms of the distance centrality of *u* and *v* in 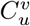 and 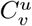 respectively. We have

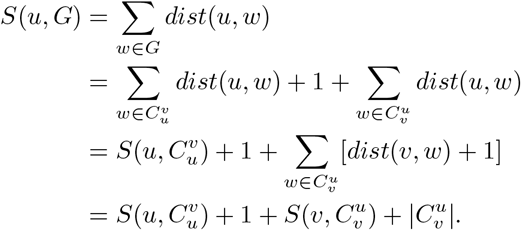

By following the same approach, we have

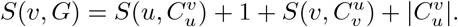

Combining two results above, we have

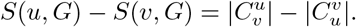

###### Lemma 5.

Let *G*^*′*^ be the pruned graph and *v ∈G \G*^*′*^. Then, *v* is either a degree-1 vertex or an endpoint of a bridge. *Proof*. Assume that *v* is not an endpoint of a bridge. Then, *v* must be contained in a cycle of *G*. The degree of *v* must be larger than or equal to 2 during pruning, which implies that *v* ∈ *G*^*′*^ and contradicts the assumption that *v* ∈ *G*\*G*^*′*^.□

By Lemma 5, we conclude that *v*_*c*_ must be an endpoint of a bridge in *G*. We consider two distinct cases based on the distance between *v*_*c*_ and *G*^*′*^ in the following, which is defined as usual by 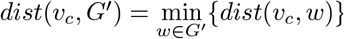.

**Case 1:** Assume that *dist*(*v*_*c*_, *G*^*′*^) = 1. Then, there is only one vertex *u ∈ G*^*′*^ such that *dist*(*v*_*c*_, *u*) = 1 and (*u, v*_*c*_) is a bridge by Lemma 5. Moreover, we have 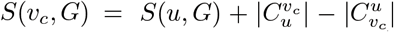 by Lemma 4. Since 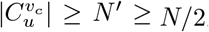 *N/*2, we have *S*(*v*_*c*_, *G*) ≥*S*(*u, G*), which contradicts the assumption that *v*_*c*_ is the distance center.

**Case 2:** Assume that *dist*(*v*_*c*_, *G*^*′*^) ≥2. From Case 1, we can deduce that for any two vertices *u* and *v* in *G G*^*′*^ and (*u, v*) is a bridge with *dist*(*u, G*^*′*^) *< dist*(*v, G*^*′*^). Then, *S*(*u, G*) *< S*(*v, G*). This implies that, for all *v* with *dist*(*v, G*^*′*^) ≥2, there is a neighbor *u* of *v* such that *S*(*u, G*) *< S*(*v, G*), which is a contradiction.

Therefore, we conclude that if *N* ^*′*^ ≥*N/*2, then *G*^*′*^ must contain the distance center.

Now, let us consider the case that *C*_*dist*_(*G*) *∉G*^*′*^. Note that for each pruned neighbor *u* of *v*^*′*^ ∈ *G*^*′*^, we know that (*v*^*′*^, *u*) is a bridge of *G*. Hence, we can compute *S*(*u, G*) immediately by using Lemma 4. In particular, since *v*^*′*^ has the minimum distance centrality on *G*^*′*^, there is a unique pruned neighbor of *v*^*′*^, say *u*^*′*^, such that *S*(*u*^*′*^, *G*) *< S*(*v*^*′*^, *G*). Since 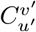 is a tree, we can apply the result of Theorem 3 in [23] to complete the rest of the proof.

### Appendix B

#### Pseudocodes

##### B.1 Triangle Motif Counting

The triangle motif counting algorithm (i.e., the computing in the feature engineering step) is implemented by Algorithm 1, where *l* is the number of disconnected components after graph pruning, *maxC* is the total number of clusters in *G*_*i*_, and MEGA(*G*_*i,j*_) is the recursive function of the feature engineering step for triangle motif counting in *G*_*i,j*_.

###### Algorithm 1: Triangle Motif Counting

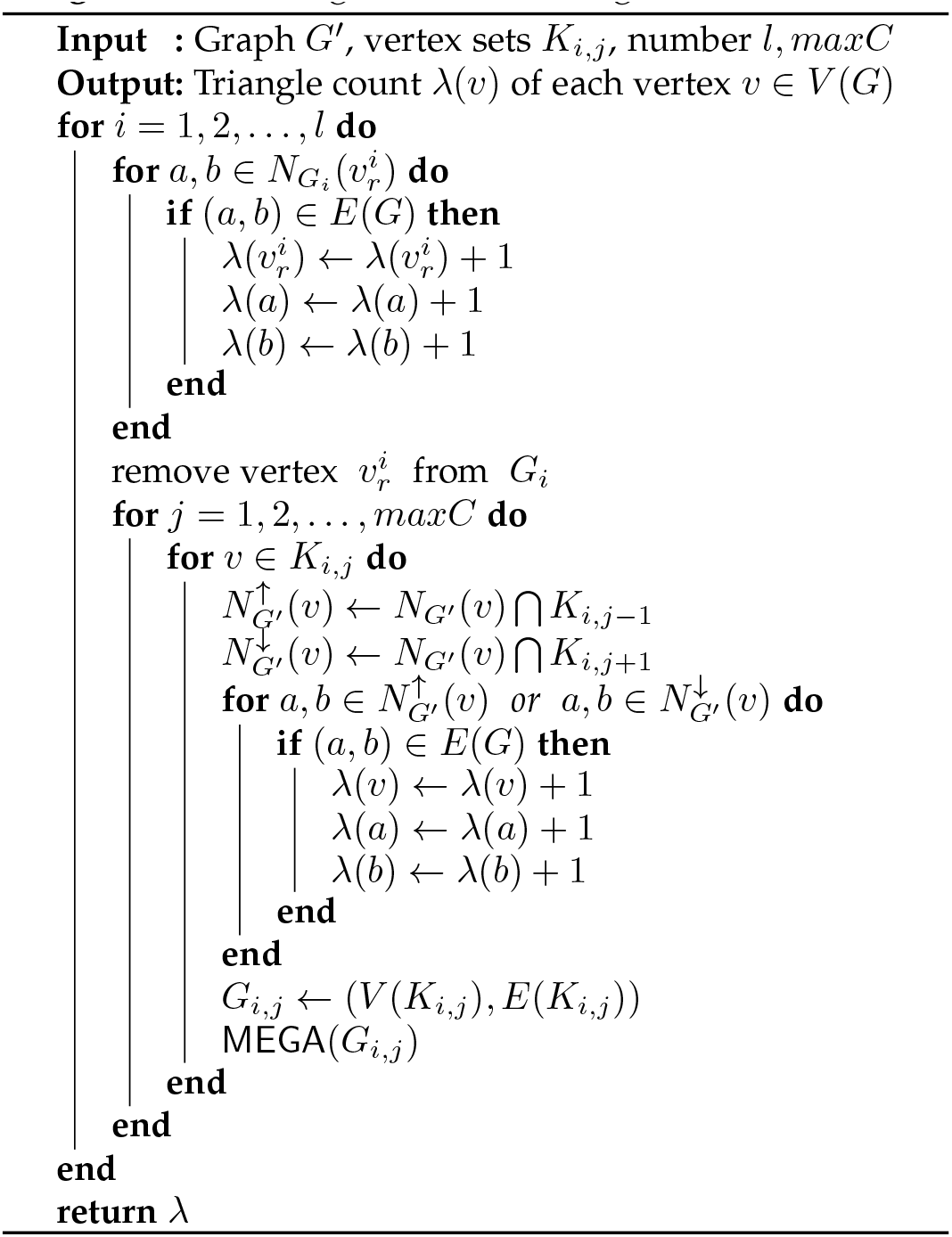

##### B.2 Distance Center Computation

Algorithm 2 describes the feature engineering step of MEGA for distance center computation in the input graph *G*. Based on Theorem 3, if *N* ^*′*^ *< N/*2, where *N* ^*′*^ is the cardinality of *V* (*G*^*′*^) and *N* is the cardinality of *V* (*G*), we can use Corollary 1 to backtrack the removed distance center in *G*.

###### Algorithm 2: Distance Center Computation

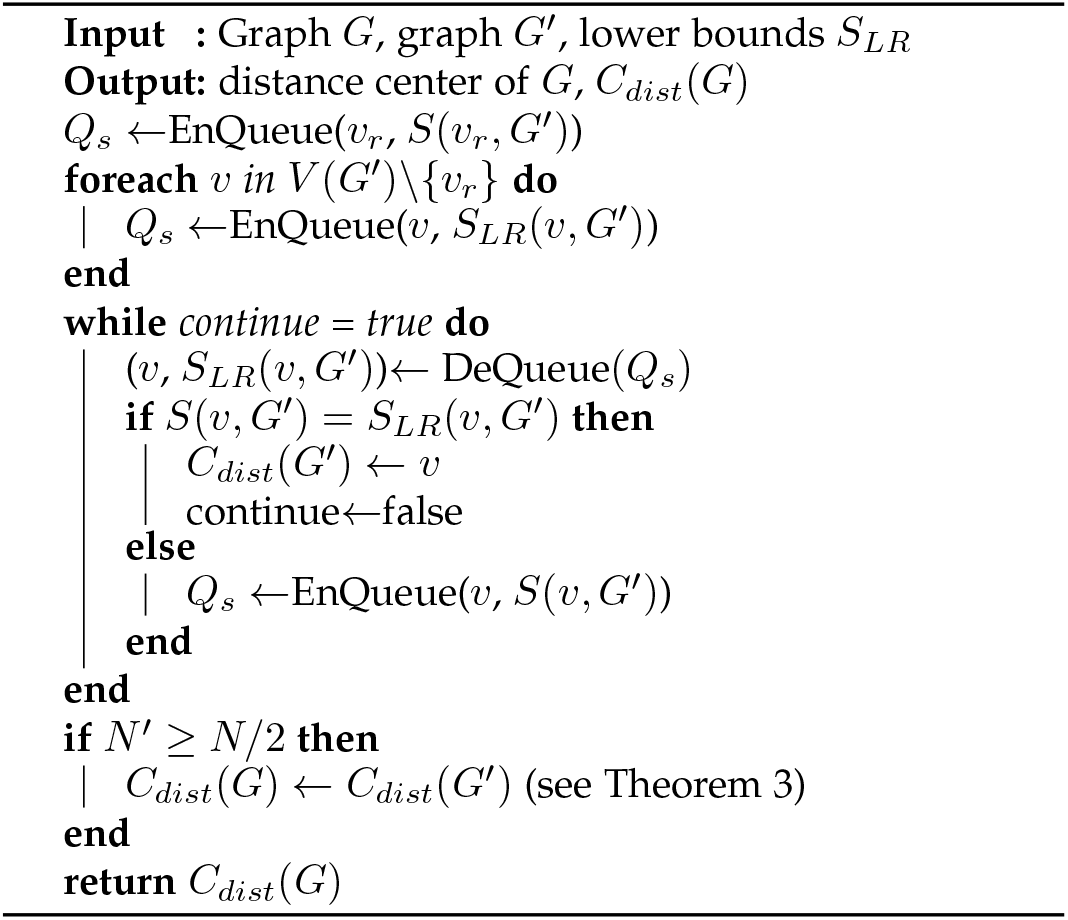

### Appendix C

#### Time Complexity OF Triangle Motif Counting

The time complexity of the graph pruning step depends on the number of pairs of vertices that we need to verify (whether an edge exists between them). Let *P* (*G*) be the set of pruned vertices of *G* with threshold *θ*, i.e., *P* (*G*) ={*v* ∈ *V* (*G*) | *v* ∈*∉ V* (*G*^*′*^)}. Then, the time complexity of the pruning step is bounded above by 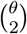 · |*P* (*G*)|. Assume that *θ* ≪ *N*. Then, the time complexity of the pruning step is bounded above by *O*(*N*) since |*P* (*G*)| ≤ *N*.

In hierarchical clustering, we select a vertex 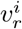 as an optimal root of the BFS for *G*_*i*_ to obtain the distances from 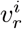 to any other vertex in *G*_*i*_. Hence, the time complexity of the hierarchical clustering step is *O*(|*V* (*G*^*′*^)| + |*E*(*G*^*′*^)|) = *O*(| *E*(*G*^*′*^)|) since we only consider graphs with |*V* (*G*^*′*^)| ≤ |*E*(*G*^*′*^) |in the triangle motif counting problem.

In the computing step, each vertex *v* has three types of neighbors: the neighbors in the upper cluster 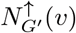, the neighbors in the lower cluster 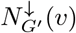, and the neighbors in the same cluster 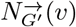. Therefore, the number of pairs of vertices that we need to check for each vertex *v* is at most

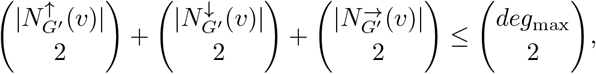

where *deg*_max_ is the maximum degree of *G*^*′*^. Thus, the time complexity of the computing step is 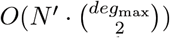.

We thus conclude that the total time complexity of the feature engineering step of MEGA for solving the triangle motif counting problem is 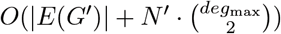.

### Appendix D

#### Illustrative Example of Distance Center Computation

We use an example to illustrate the difference of finding the message source estimator between the feature engineering step of MEGA and the BFS heuristic algorithm in [9]. Using the same example as in [9], we have a spread graph as shown in Fig. 14. There are 5 vertices received the message (*v*_1_ to *v*_5_), and we want to find the distance-based message source estimator of this spread graph using MEGA.

The input graph has five vertices (*v*_1_ to *v*_5_) that received the message, and our goal is to find the distance-based message source estimator using the MEGA framework. In the first step of graph pruning with *θ* = 1, we remove *v*_5_ and update its parent *v*_4_ such that *T* (*v*_4_) = 2 and *D*(*v*_4_) = 1, resulting in the pruned graph *G*^*′*^. We then use every vertex in the pruned graph as the root to perform BFSs and determine the worst-case performance. Starting with *v*_1_ as the root, we obtain the following results:

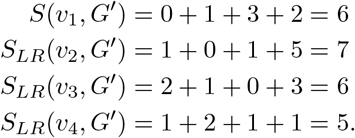

Since *S*_*LR*_(*v*_4_, *G*^*′*^) is the smallest, we need to check if *S*(*v*_4_, *G*^*′*^) = *S*_*LR*_(*v*_4_, *G*^*′*^) by using *v*_4_ as the root for the second BFS tree traversal. As we have *S*(*v*_4_, *G*^*′*^) = *S*_*LR*_(*v*_4_, *G*^*′*^) and the lower bound of every vertex remains unchanged, we find that *v*_4_ is the distance-based message source estimator of *G*^*′*^. Since *N* ^*′*^ *> N/*2, based on Theorem 3, we conclude that *v*_4_ is the message source of *G*.

Similarly, for the roots *v*_2_ and *v*_3_, we need to perform two BFS tree traversals as well. However, for the root *v*_4_, we have

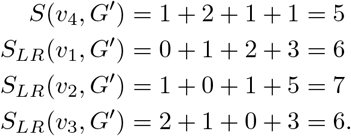

Since *S*(*v*_4_, *G*^*′*^) is already the smallest, we conclude that *v*_4_ is the message source of *G* by using only one BFS tree. As such, in the worst case, we only need to perform BFS twice for finding the distance-based message source estimator of the spread graph. Meanwhile, *v*_4_ is also identified as the rumor center using the BFS heuristic algorithm in [9] but it requires to construct BFS trees starting from every vertex in the spread graph. Thus, in this example, the number of BFSs performed and the number of edges visited by MEGA are both less than that of the algorithm in [9].

### Appendix E

#### Experimental Performance Evaluation

In this section, we provide more detailed experiments to assess the performance of the feature engineering step of MEGA for triangle motif counting and distance center computation on (simple undirected) social networks.

##### E.1 Triangle Motif Counting on Social Networks

We evaluate the performance of the feature engineering step of MEGA on the triangle motif counting problem using the social networks provided by Stanford Large Network Dataset Collection (SNAP) [44].

###### E.1.1 Optimal Threshold Tuning

Fig. 15 shows how the threshold *θ* affects the computation time of the feature engineering step of MEGA for triangle motif counting on eight real-world networks provided by SNAP. We also compare the performance of MEGA using the degree center and a random vertex as the BFS root in hierarchical clustering since other centrality measures such as PageRank require global information from all other vertices that must increase the computational complexity. We see that MEGA with the degree center performs better than that with a random vertex. We also note that finding an optimal threshold for an arbitrary graph is not a straight-forward task unless we further exploit the inherent structure of the graph.

**Fig. 15.**
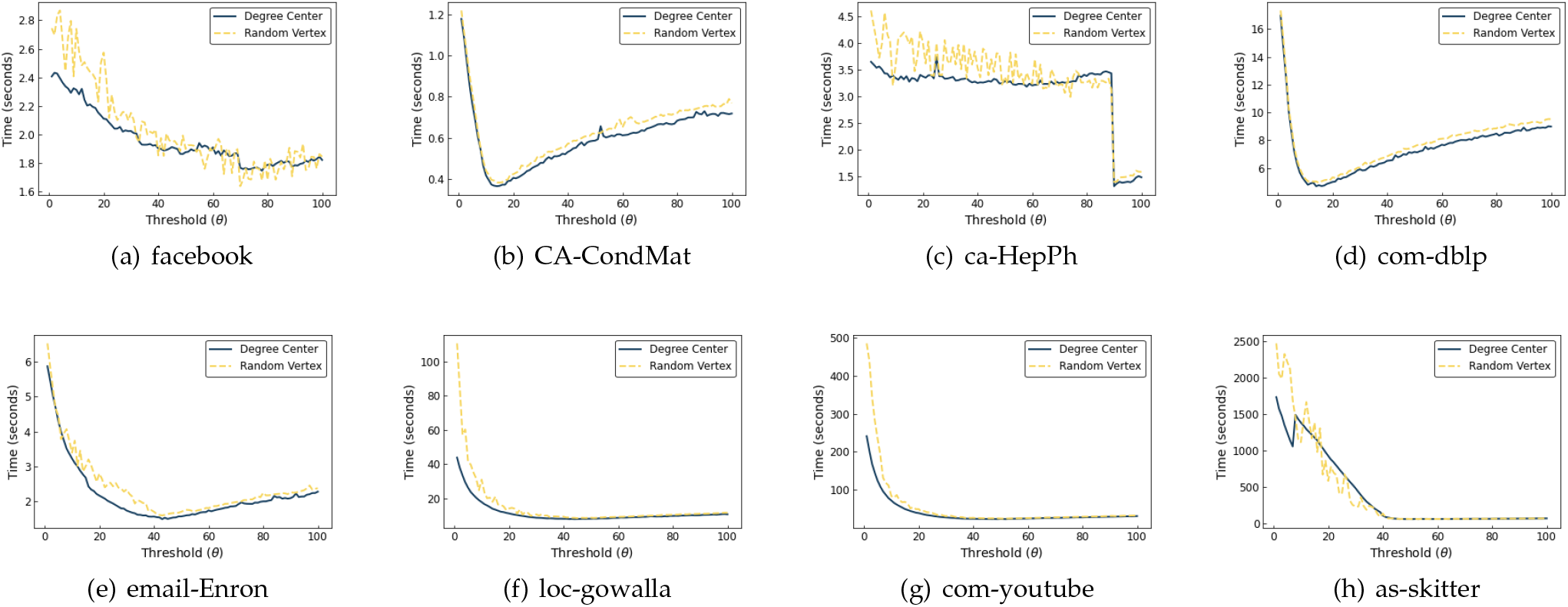
The performance of MEGA on eight real-world networks with different thresholds *θ* ranging from 0 to 100. The y-axis is the running time in seconds, and the x-axis is the value of *θ*. We also compare the performance of MEGA using the degree center (blue) and a random vertex (yellow) as the BFS root for hierarchical clustering.

Based on Lemma 1, we use a data-driven (statistical) model to approximate the optimal threshold *θ*^∗^ for MEGA. We generate 6,000 synthetic networks in which the optimal thresholds *θ*^∗^ are computed. Each network has 1,000 vertices, and the number of edges of each network is not fixed. Note that the number of vertices of the input graph is not a critical parameter for computing *θ*^∗^. In Fig. 16, we see that the gradient of the blue line, which is a linear regression model that fits the results, is relatively small that keeps *θ*^∗^ always less than 100, even the graph size starts to increase. We also note that noisy data only appear when the graph size is comparatively small. Therefore, the optimal threshold *θ*^∗^ computed by this model must satisfy the condition of *θ*^∗^ ≪ *N* and balance the trade-off for large-scale networks.

**Fig. 16.**
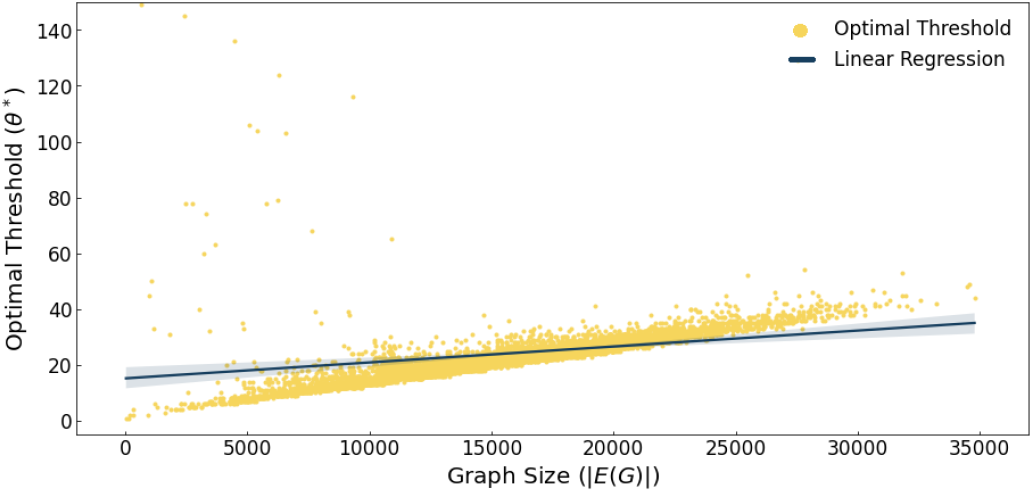
Optimal threshold tuning on 6,000 synthetic networks based on Lemma 1. The y-axis is the value of the optimal threshold *θ*^∗^, and the x-axis is the graph size |*E*(*G*) | of each network. The blue line is a linear regression model that fits the results.

###### E.1.2 Evaluation on Social Networks

We compare MEGA with the algorithm in [45], which is an award-winning work of the MIT/Amazon/IEEE Graph Challenge. The algorithm in [45] assigns direction to each edge based on the degree of each vertex. It implies that, for each vertex *v*, there are 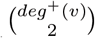pairs of vertices needed to be checked, where *deg*^+^(*v*) ^2^ is the outdegree of *v*. Hence, its time complexity is 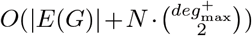, where |*E*(*G*)| is the time complexity of the direction assignment and 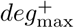 is the maximum outdegree. This time complexity is the same as that of MEGA (cf. Appendix C), and this is why we choose it as a baseline. In Fig. 17, we see that MEGA with *θ*^∗^ outperforms the algorithm in [45] on different social networks by averagely 25.3 times faster.

**Fig. 17.**
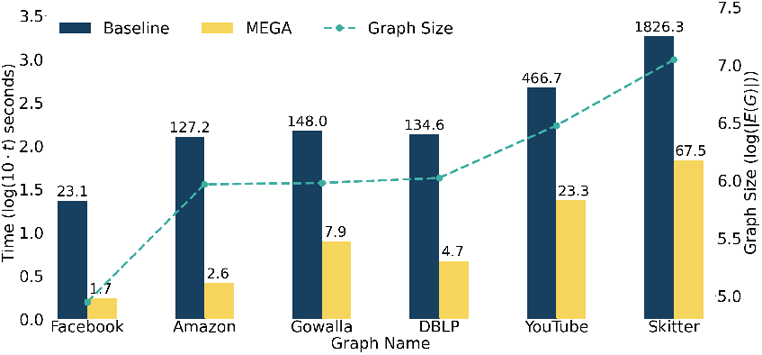
Comparison between MEGA and the algorithm in [45] (which we set as baseline) on six social networks. The primary y-axis (blue) is the running time *t* in seconds, and the secondary y-axis (green) is the graph size |*E*(*G*) |. A log scale is used for these two axes, and we multiply *t* by 10 to avoid negative values (i.e., primary = log(10 *t*) seconds and secondary = log(|*E*(*G*) |)). The number on top of each bar is the exact running time of both algorithms.

###### E.1.3 Evaluation on Synthetic Random Networks

We compare the performances of MEGA and the algorithm in [45] on ten large-scale synthetic random networks generated by the BA model and the ER model respectively.

For BA network, we randomly generate the two parameters (*m*_0_, *m*) for the BA model so as to obtain ten random networks, where each network contains 1 million vertices and 9, 999, 945 edges. We use Theorem 1 to compute the optimal threshold *θ*^∗^ for each BA network such that MEGA can optimally decompose the BA networks in pruning. In Fig. 18(a), we see that although some vertices have a large degree, MEGA can still be able to decompose the networks with *θ*^∗^ and outperform the algorithm in [45]. Moreover, since we only execute the pruning step with a small threshold (*θ*^∗^ ≪ *N*), the time complexity of MEGA is in *O*(*N*).

**Fig. 18.**
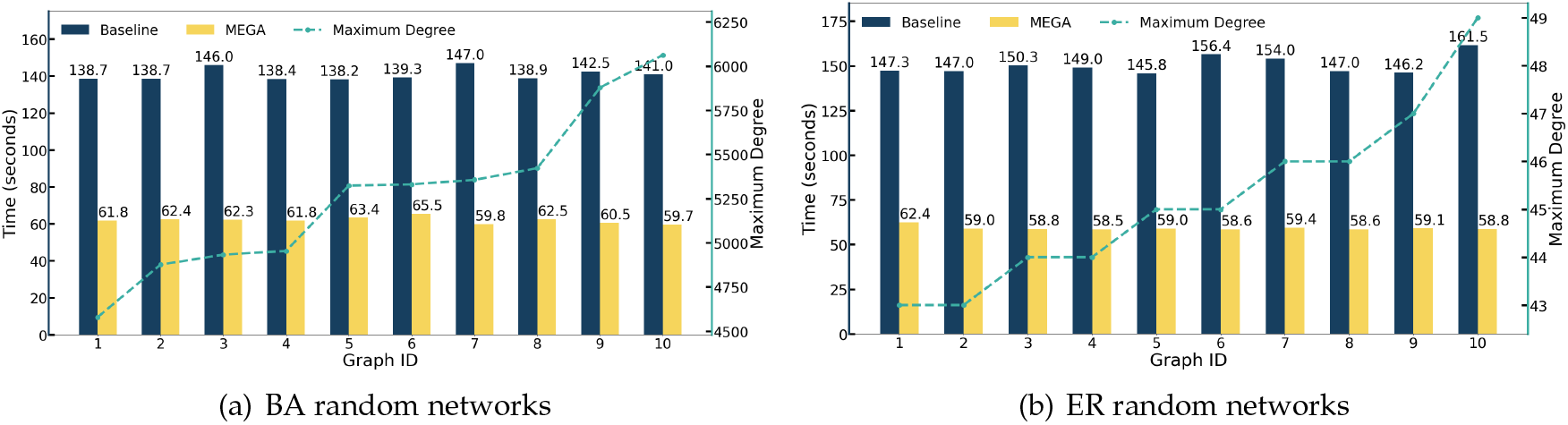
Comparison between MEGA and the algorithm in [45] (which we set as baseline) on ten BA random networks (left) and ten ER random networks (right) respectively. Each random network has 1M vertices and 9.9M edges. The primary y-axis (blue) is the running time in seconds, and the secondary y-axis (yellow) is the maximum degree of each random network.

Similarly, we generate ten ER random networks with the same size as the BA networks. In Fig. 18(b), we observe that MEGA beats the algorithm in [45] with *θ*^∗^. Note that the probability of a given vertex having degree greater than *θ*^∗^ in the ER networks is less than 8%, which implies that the ER networks are largely decomposed in pruning.

##### E.2 Distance Center Computation on Social Networks

In this section, we assess the performance of the feature engineering step of MEGA for distance center computation on different social networks provided by SNAP [44].

We consider both performance improvement and accuracy on single message source detection. We first compare MEGA with the algorithm presented in [29] based on the speedup which has been defined in Section 7.4.1 and then compare the accuracy of single message source detection with the work in [9].

###### E.2.1 Evaluation on Social Networks

The graph pruning of our MEGA performs very well on graphs with large diameter, outperforming the NBBound proposed in [29]. However, the efficacy of graph pruning in small-world networks is relatively low since the graph diameter of such networks is small. To further increase the speedup of MEGA, we utilize the *Multi-Source BFS (MS-BFS)* [46]. The idea is motivated by the observation that, when running a large number of BFSs sequentially, most of the edges are visited multiple times, which might deteriorate the overall performance. The MS-BFS algorithm addresses this problem by running BFSs from multiple sources concurrently. When a set of BFSs visits the same vertex, this vertex will be visited only once, and its information will be shared with all BFSs in the set. Thus, a large number of visits can be shared by multiple BFSs when applying the MS-BFS algorithm. In Fig. 19, we see that MS-BFS greatly increases the speedup of MEGA.

**Fig. 19.**
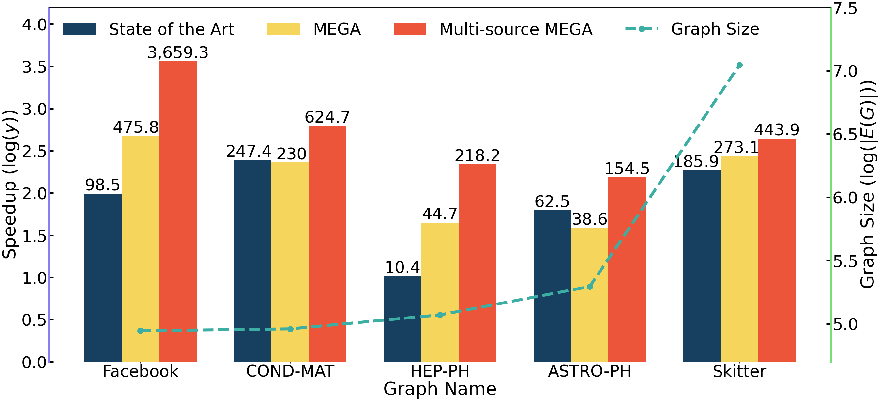
Comparison between MEGA and the algorithm in [29] (which we set as baseline) on five social networks. The primary y-axis (blue) is the speedup *y*, and the secondary y-axis (green) is the graph size |*E*(*G*) |. A log scale is used for these two axes. The number on top of each bar is the exact speedup of both algorithms.

###### E.2.2 Single Message Source Detection in Social Networks

We perform simulations on six social networks and compare the performances of our MEGA framework with the BFS heuristic in [9]. For each graph, we randomly select a message source vertex and let the message spread to 100 vertices. We then apply MEGA and the BFS heuristic in [9] respectively to find a source estimator. Note that the error indicates the graph distance from the source estimator to the real source vertex. We perform over 500 simulations and calculate the average error and speedup for each graph. In Fig. 20, we can observe that the speedup of our MEGA framework is significantly larger than that of the BFS heuristic in [9] and there is only a slight difference in accuracy (average error) between these two approaches.

**Fig. 20.**
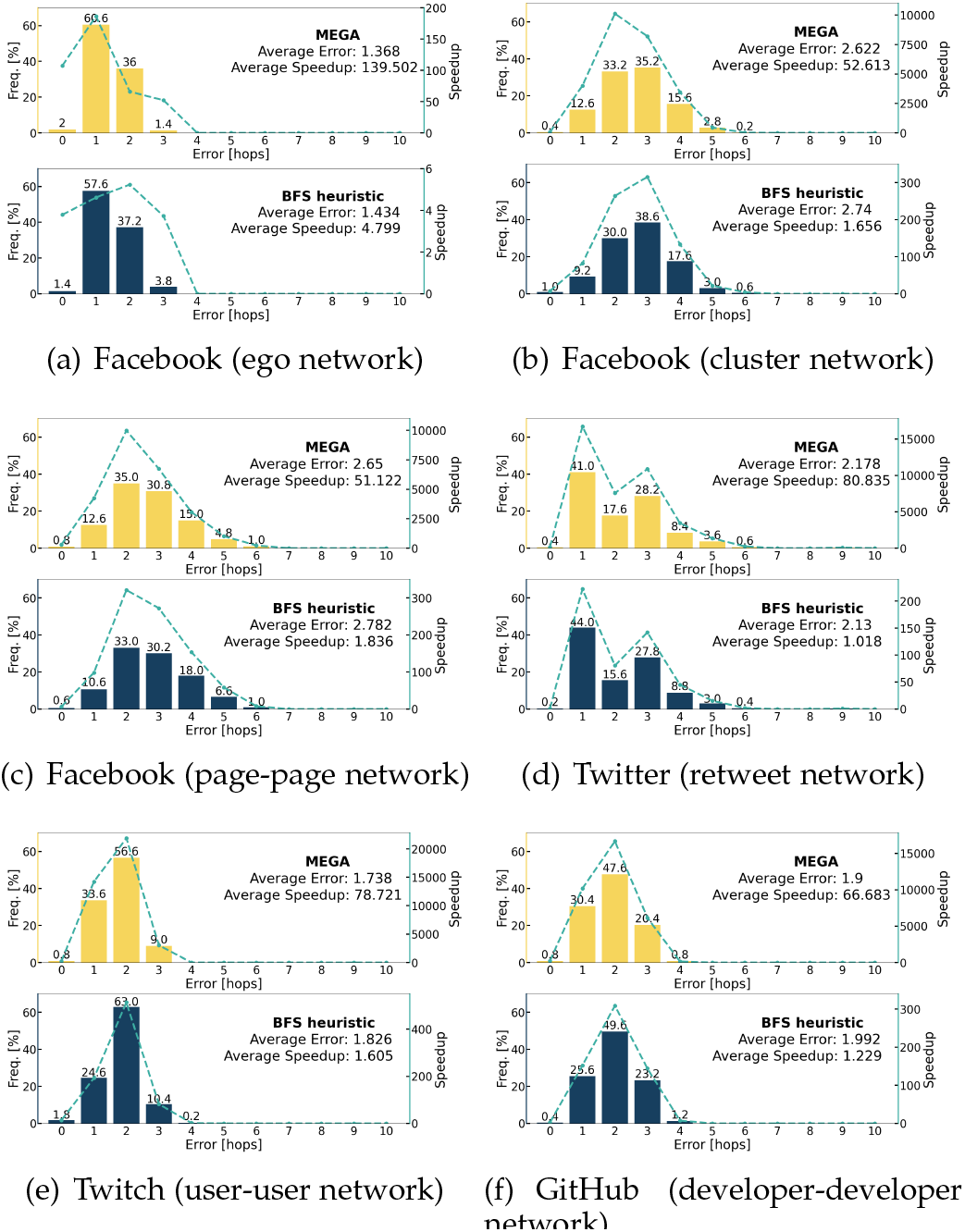
Histograms of the error for MEGA (yellow) and the BFS heuristic in [9] (blue) on six different types of social networks with 100 vertices received the message in the spread graph. The green dotted line is the average speedup for each error and the average error is the average distance (in terms of hop count) between the real source and the estimators calculated by both methods over 500 simulations.

**Fig. 21.**
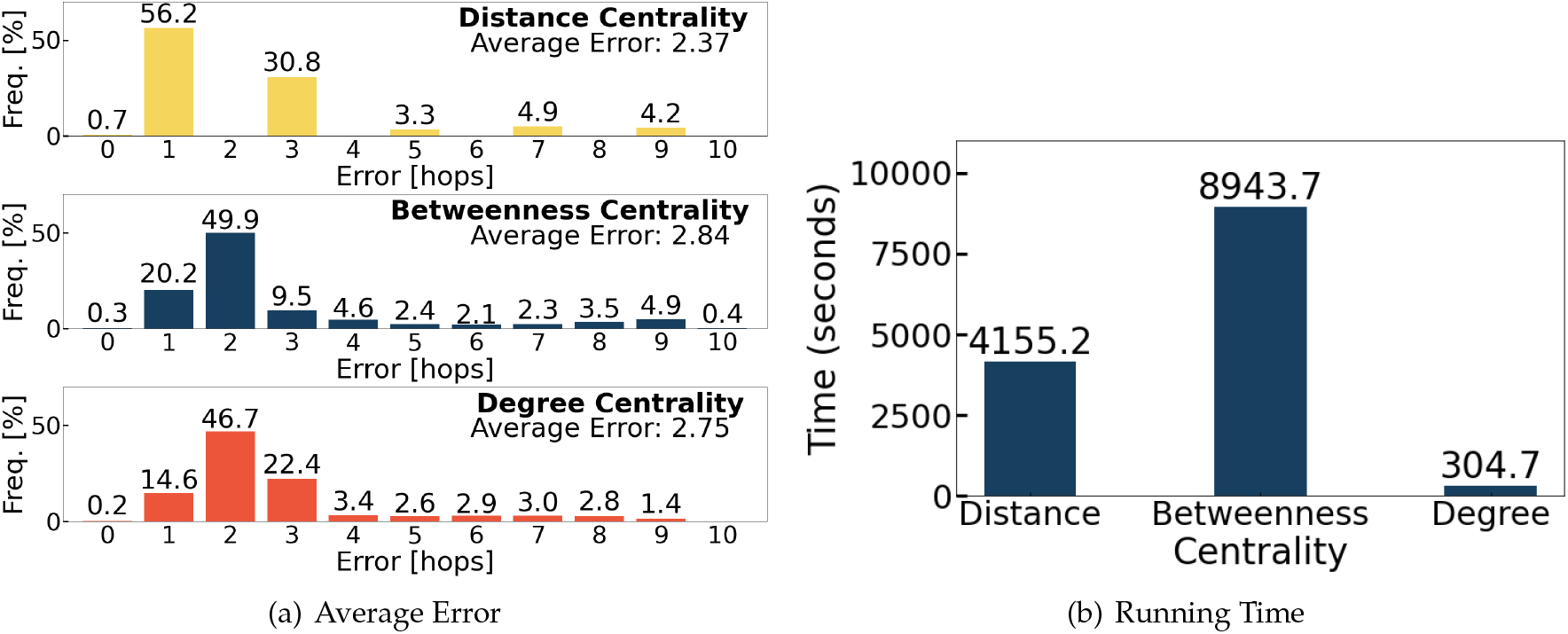
Comparison of the average error and running time (in seconds) in identifying prominent vertices in the retweet network using distance centrality (MEGA), betweenness centrality, and degree centrality as the metrics.

### Appendix F

#### Small-world Analysis

We have identified three parts in which finding the most prominent vertices of the graph is required: (1) finding the root for BFS in hierarchical clustering for triangle counting, finding the root for BFS in hierarchical clustering for distance center computation, and (3) using distance centrality to find influential vertices. To address (1), we suggest using the degree center as the root, as it does not require additional complex computation. In contrast, other approaches could lead to increased computational complexity, and choosing a random root would be meaningless. By choosing the degree center as the root, it’s possible to minimize the total number of clusters and potentially reduce computational complexity. However, further analysis is required for (2) and (3), which we will discuss below.

To perform our analysis, we generate five small-world networks using the Watts-Strogatz model, with each network containing 50, 100, 500, 1,000, and 2,000 vertices, respectively. In each network, each vertex is connected to 60% of its nearest neighbors in a ring topology, with each edge having a rewiring probability of 0.5. We identify hubs as vertices with high degree, low distance centrality, or high betweenness centrality. The number of edges for each network is listed below:

| 50 Vertices | 100 Vertices | 500 Vertices | 1,000 Vertices | 2,000 Vertices |
| --- | --- | --- | --- | --- |
| 750 | 3,000 | 75,000 | 300,000 | 1,200,000 |

For (2), we suggest selecting a vertex *v* with minimum degree in the pruned graph, followed by selecting a vertex with the largest eccentricity (i.e., maximum distance away from *v*) as the root. We compare the performance of this approach with using the degree center as the root, as it is not appropriate to use hubs with low distance centrality or high betweenness centrality as the root in this step when our main objective is to identify the distance center. The results below indicate that for small-world networks, using the degree center can reduce the algorithm’s running time, even though the speedups of both approaches are the same. As the majority of vertices have low distance centrality and high degree centrality, most neighboring vertices can be reached by a small number of hops. Therefore, the speedups for both approaches should be almost equivalent. Additionally, the efficacy of the graph pruning step of the MEGA framework in small-world networks is relatively low since the graph diameter of such networks is small, and the graph pruning is not triggered. As a result, the framework’s average performance on small-world networks is only moderate, although the speedups appear relatively large, they are only proportional to the number of edges in the graphs. However, in the retweet network, our proposed approach outperforms the small-world analysis approach. Therefore, we agree that if the input graph exhibits small-world network properties, the small-world analysis approach can provide better performance in terms of running time. Still, if the input is unlikely to be a small-world network, our proposed approach is more efficient.

For (3), when dealing with small-world networks that have more than 100 vertices, we randomly choose a message source vertex and allow the message to propagate to 100 vertices (at most) for each message spread graph. For small-world networks with 100 vertices or less, we randomly choose a message source vertex and allow the message to propagate to the entire graph. To find the source estimator, we utilize distance centrality (low), degree centrality (high), and betweenness centrality (high). For small-world networks, we use MEGA to compute distance centrality to find the distance center, Stanford Network Analysis Platform (SNAP) to compute betweenness centrality to find the betweenness center and SNAP to find the degree center. In the retweet network, we use the same methods used in small-world networks to identify the top 5,195 vertices with the smallest distance centrality, largest betweenness centrality, and largest degree centrality, respectively. The error indicates the graph distance from the source estimator to the real source vertex. We conduct over 500 simulations and calculate the average error and average running time for each centrality measure in each small-world network. The results below show that in small-world networks, the average errors for the hubs with high degree and high betweenness centrality are slightly higher than those for distance centrality. Additionally, in the retweet network, the performance of using distance centrality is better than that of degree and betweenness centrality. While using small-world analysis could potentially decrease computational costs since degree centrality is the fastest in finding the hubs, betweenness and distance centrality require much longer time, especially for betweenness centrality. The trade-off is that the accuracy will be lower if such a faster approach is applied. Therefore, we choose distance centrality as it is more accurate, but we also optimize the process of finding hubs with low distance centrality to balance the trade-off.

## Notes

This research was supported in part by the National Science and Technology Council, R.O.C., under Grant 112-2115-M-033-004-MY2, the Ministry of Education, Singapore, under its Academic Research Fund (No. 022307 and AcRF RG91/22), a grant from the NTU World Health Organization Collabo-rating Centre for Digital Health and Health Education, and a grant from the Hong Kong ITF Project No. ITS/188/20.

### Competing Interest Statement

The authors have declared no competing interest.

### Summary of Updates

Fixed typos in Section 3 and added paper info in P.1

